# SARS-CoV-2 lineage dynamics in England from January to March 2021 inferred from representative community samples

**DOI:** 10.1101/2021.05.08.21256867

**Authors:** Oliver Eales, Andrew J. Page, Sonja N. Tang, Caroline E. Walters, Haowei Wang, David Haw, Alexander J. Trotter, Thanh Le Viet, Ebenezer Foster-Nyarko, Sophie Prosolek, Christina Atchison, Deborah Ashby, Graham Cooke, Wendy Barclay, Christl A. Donnelly, Justin O’Grady, Erik Volz, The COVID-19 Genomics UK (COG-UK) Consortium, Ara Darzi, Helen Ward, Paul Elliott, Steven Riley

## Abstract

Genomic surveillance for SARS-CoV-2 lineages informs our understanding of possible future changes in transmissibility and vaccine efficacy. However, small changes in the frequency of one lineage over another are often difficult to interpret because surveillance samples are obtained from a variety of sources. Here, we describe lineage dynamics and phylogenetic relationships using sequences obtained from a random community sample who provided a throat and nose swab for rt-PCR during the first three months of 2021 as part of the REal-time Assessment of Community Transmission-1 (REACT-1) study. Overall, diversity decreased during the first quarter of 2021, with the B.1.1.7 lineage (first identified in Kent) predominant, driven by a 0.3 unit higher reproduction number over the prior wild type. During January, positive samples were more likely B.1.1.7 in younger and middle-aged adults (aged 18 to 54) than in other age groups. Although individuals infected with the B.1.1.7 lineage were no more likely to report one or more classic COVID-19 symptoms compared to those infected with wild type, they were more likely to be antibody positive 6 weeks after infection. Viral load was higher in B.1.1.7 infection as measured by cycle threshold (Ct) values, but did not account for the increased rate of testing positive for antibodies. The presence of infections with non-imported B.1.351 lineage (first identified in South Africa) during January, but not during February or March, suggests initial establishment in the community followed by fade-out. However, this occurred during a period of stringent social distancing and targeted public health interventions and does not immediately imply similar lineages could not become established in the future. Sequence data from representative community surveys such as REACT-1 can augment routine genomic surveillance.

## Introduction

Since the emergence of SARS-CoV-2 in late 2019 [1] there has been a continuous accumulation of mutations leading to a genetically diverse phylogeny [2]. Although most mutations are neutral, having no effect on the epidemiology of the virus, some have been found to affect transmissibility [3] and antigenicity [4], and have arisen on multiple occasions in independent lineages [5]. Lineages that are judged likely to have increased transmissibility or severity relative to current dominant lineages in the UK are termed ‘Variants of Concern’ (VOC) [6–8].

The B.1.1.7 lineage (Pango nomenclature [9], used for the rest of the paper) VOC was first detected in Kent, England 20th September 2020 [10,11]. Since its emergence, it has risen to become the dominant lineage in the United Kingdom (UK), and has increased in frequency in many other countries [12]. Previous studies have estimated that this lineage is more transmissible than previously dominant lineages, as measured by the reproduction number (R) [6,13].

The B.1.351 lineage was first detected in South Africa [7] in October 2020 and by March 2021 there had been 291 detections in the UK [14]. The lineage is associated with the E484K single nucleotide polymorphism (SNP) in the spike protein, which has been found to reduce the neutralizing activity of post-vaccination sera [15], triggering fears of lowered vaccine efficacy. This SNP has also been detected in a cluster of B.1.1.7 cases In England, predominantly in the South West [16]. Two further lineages, A.23.1 [17] and B.1.525, are currently described as ‘Variants under Investigation (VUI)’ [18] due to the presence of several SNPs of biological significance. Both of these lineages have been detected in low numbers in the United Kingdom [19,20]. A cluster of A.23.1 that exhibits the E484K SNP has been detected in Liverpool, England [16].

Extensive genomic surveillance has been undertaken in the UK by the COVID-19 Genomics UK Consortium (COG-UK) [21]. From its inception in March 2020 to the end of March 2021, COG-UK sequenced over 320,000 positive cases [22] representing 45% [23] of all uploaded sequences to GISAID, a global open-access database for coronavirus and influenza genomic data [24], with UK coverage of all detected samples varying from 2.5% [25] to 17.1% [26]. Samples included in COG-UK data are taken from several different sources: hospital cases, routine community surveillance, outbreak investigations and border screening.

The REal-time Assessment of Community Transmission study obtains throat and nose swabs from a random sample of the population in England (REACT-1) [27]. From the beginning of May 2020 to the end of March 2021, there were 10 rounds of REACT-1 with between 140,000 and 175,000 swab tests each round. Here we present the results of the genome sequencing performed on the positive swabs in early 2021, from round 8 (6 January - 22 January), round 9 (4 February - 23 February) and round 10 (11 March −30 March).

## Results

### Lineage diversity

In round 8 we were able to reliably determine lineages for 1,088 out of 2,282 positive samples, of which 83% (80%, 85%, n=898) were the B.1.1.7 lineage, 0.37% (0.14%, 0.94%, n=4) were the B.1.351 lineage, 0.18% (0.05%, 0.67%, n = 2) were the A.23.1 lineage, 0.09% (0.02%, 0.52%, n=1) were the B.1.525 lineage, and 0.18% (0.05%, 0.67%, n=2) were B.1.1.7 lineage with the E484K SNP (B.1.1.7+E484K, first identified in a cluster of cases in Bristol, UK) (Supplementary Table 1). The remaining 17% (15%, 19%, n=181) of lineages were classified as wildtype and comprised 35 distinct lineages, the main constituent of which was B.1.177 (n=105). In round 9, 236 lineages out of 689 positives were determined, of which 96% (92%, 98%, n=226) were the B.1.1.7 lineage and the remaining 4.2% (2.3%, 7.6%, n=10) were classified as wildtype. In round 10, all 73 lineages determined from 227 positive samples were B.1.1.7. Despite the reduced number of samples in round 10, the data across all three rounds securely describe a decrease in diversity (*P* < 0.001 for reduction in proportion not B.1.1.7).

### Quantifying transmissibility of B.1.1.7

Fitting a logistic regression model to whether a sample was identified as B.1.1.7 or not allowed us to estimate a difference in daily growth rate between B.1.1.7 and all other lineages of 0.049 (0.034, 0.067) (Figure 1). This corresponds to an additive R advantage of 0.31 (0.21, 0.42), (assuming a mean generation time of 6.29 days, see Methods) and suggests a smaller difference than estimates based on sequences collected during November and December 2020 in england (between 0.4 and 0.7) [28]. Our lower estimate of the difference in R at a later time is consistent with a decreasing selection coefficient reported in the earlier study.

**Figure 1.**
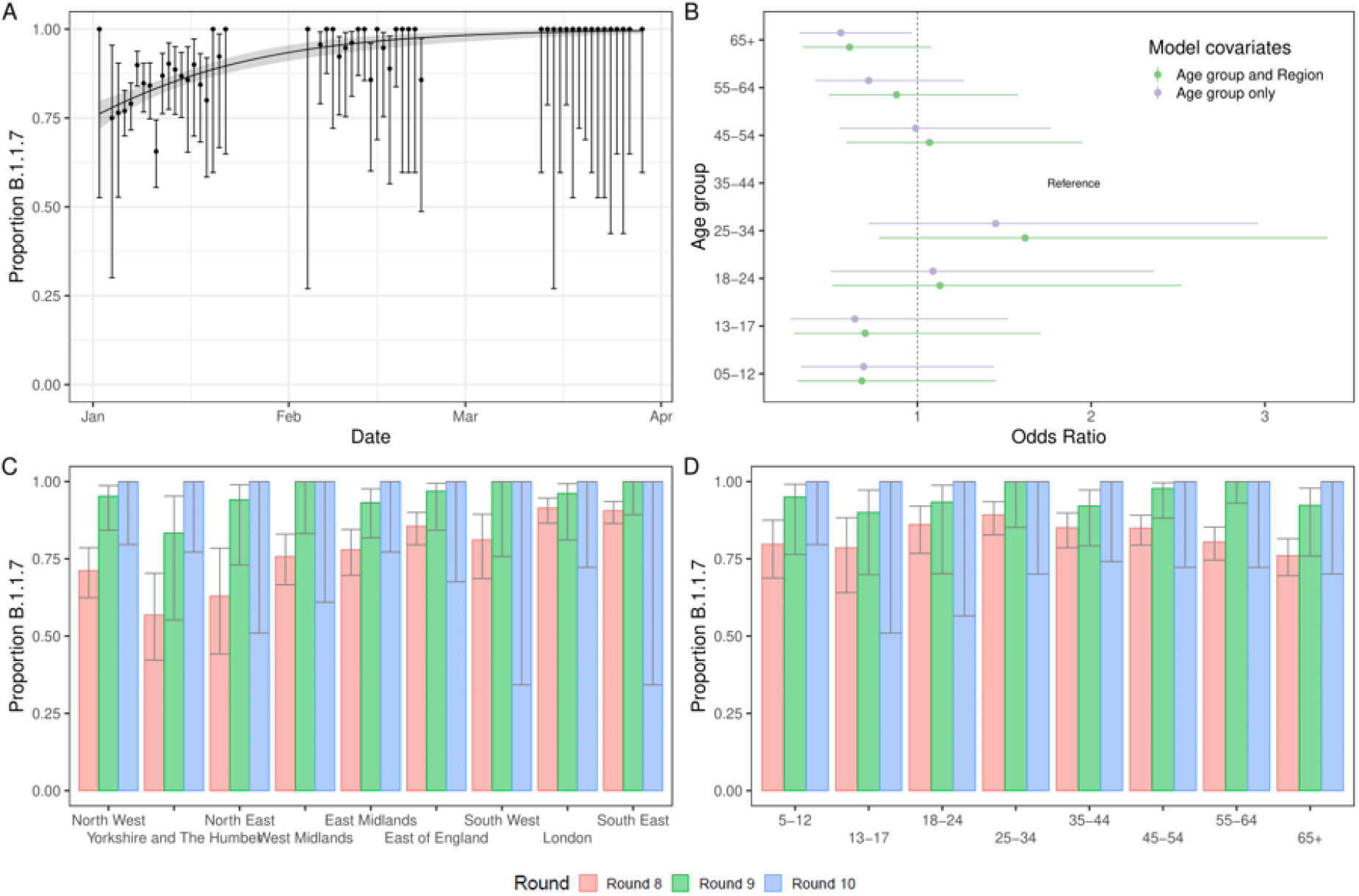
B.1.1.7 lineage in England from January to March 2021. (A) Proportion of B.1.1.7 lineage over time. Points show raw data with error bars representing the 95% confidence interval. Shaded region shows best fit Bayesian logistic regression model with 95% credible interval. (B) Odds ratio of a determined lineage being B.1.1.7 by age group for logistic models including just age group (purple) and both age group and region (green) fit to data from round 8 only. (C) Proportion of positive tests that are from the B.1.1.7 lineage by region of England. Error bars show the 95% confidence intervals. (D) Proportion of positive tests that are from the B.1.1.7 lineage by age group. Error bars show the 95% confidence intervals.

Proportions of B.1.1.7 showed marked spatial heterogeneity in January (Round 8), with regions in the Midlands and the North of England showing lower proportions of B.1.1.7 compared to regions in the South (Figure 1, Supplementary Table 2). Sub-regional analysis showed a similar trend with a smoothed term regression model (see Methods) showing lower proportions of B.1.1.7 in areas of the Midlands, Yorkshire and The Humber, and the North West (Figure 2). This pattern is consistent with prior observations of B.1.1.7 lineage emerging in the South East [11] leading to earlier seeding events in the South of England. By February (round 9), spatial heterogeneity was substantially reduced with B.1.1.7 accounting for over 80% of lineages in all regions.

**Figure 2.**
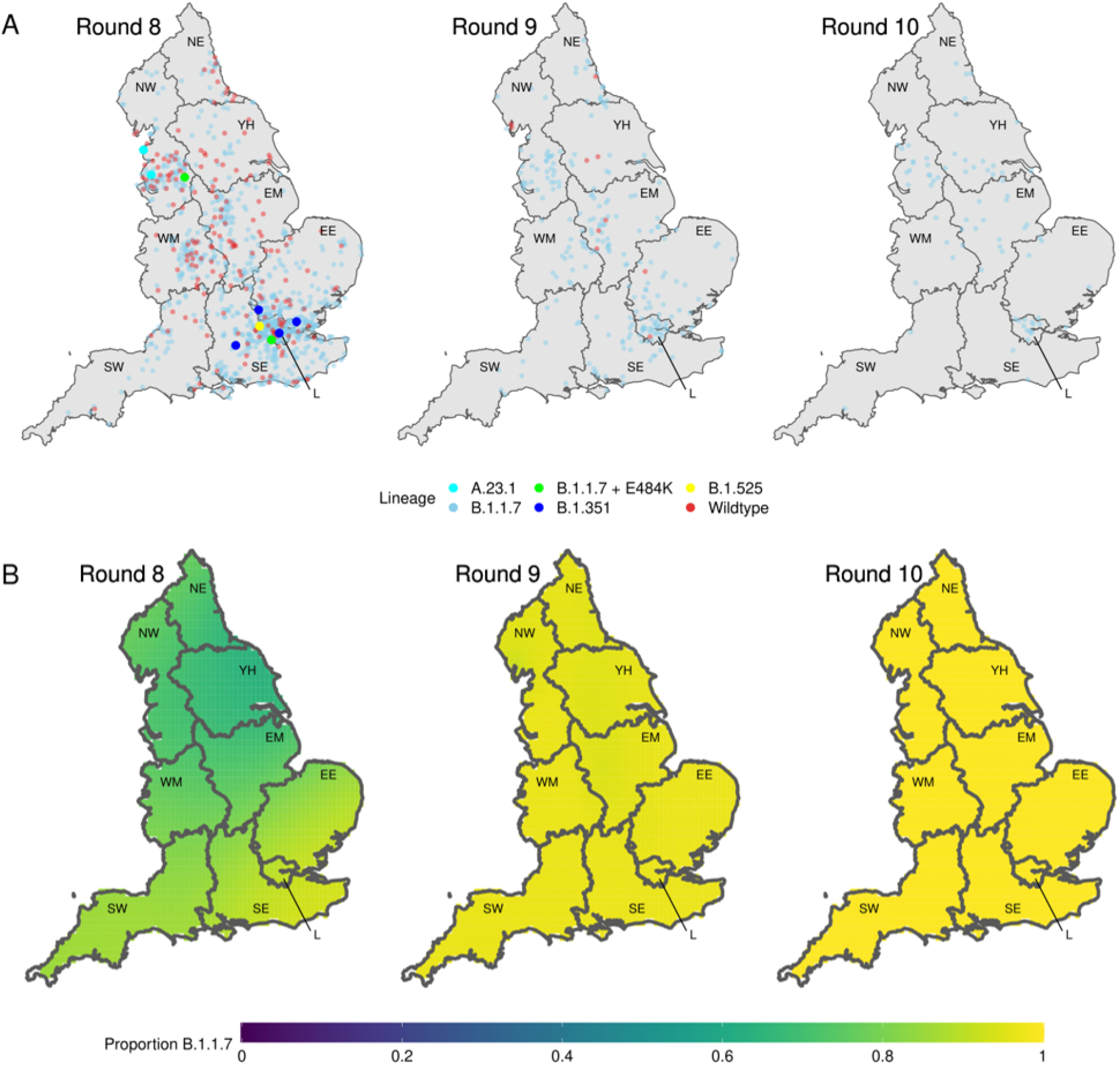
Geospatial patterns of lineage frequency. (A) Location of all positive samples for which we have identified their lineage for each round (each point moved randomly a small distance). (B) Modelled proportion of B.1.1.7 lineage across space for round 8, round 9 and round 10. Regions: NE =North East, NW = North West, YH =Yorkshire and The Humber, EM = East Midlands, WM =West Midlands, EE = East ofEngland, L = London, SE = South East, SW = South West.

During round 8 there were higher proportions of B.1.1.7 in 18-to 54-year olds compared with other age groups (Figure 1). Using a logistic regression model (see Methods), this pattern was not explained by regional confounding (Figure 1). In contrast, for round 9, albeit based on fewer positive samples, the proportion of B.1.1.7 was similar in all age groups. Case data from November and December 2020 show a higher proportion of B.1.1.7 in school aged children than in other age groups [6], however schools were open for face-to-face teaching for all children during this time unlike during our study when school attendance was greatly limited [29,30].

### Rates of symptom reporting

The percentage of people infected with B.1.1.7 reporting no symptoms (Figure 3, Supplementary Table 3) in the week prior to providing a swab was 33.3% (30.4%, 36.3%), compared with 38.2% (30.8%, 46.1%) for wildtype (*P*=0.24). Looking at the percentage of people reporting COVID-19 like symptoms (loss or change of sense of taste, loss or change of sense of smell, new persistent cough, fever) in the last week we found similar percentages exhibiting these symptoms between lineages with 45.7% (42.6%, 48.8%) for B.1.1.7, compared with 45.4% (37.7%, 53.3%) for the wildtype (*P*=0.94). Our results describe only the lower part of the severity pyramid -- the fraction of those infected who develop symptoms -- and contrast with a previous study using clinical cases as the denominator which found that the B.1.1.7 lineage caused more severe illness with increased relative mortality [31]. However, other previous work found no difference in symptomatology for B.1.1.7 against other circulating lineages [32]. We also note that participants in REACT-1 were not followed up, other than a small subset in round 8. Therefore, some participants will have developed symptoms after filling out the questionnaire.

**Figure 3.**
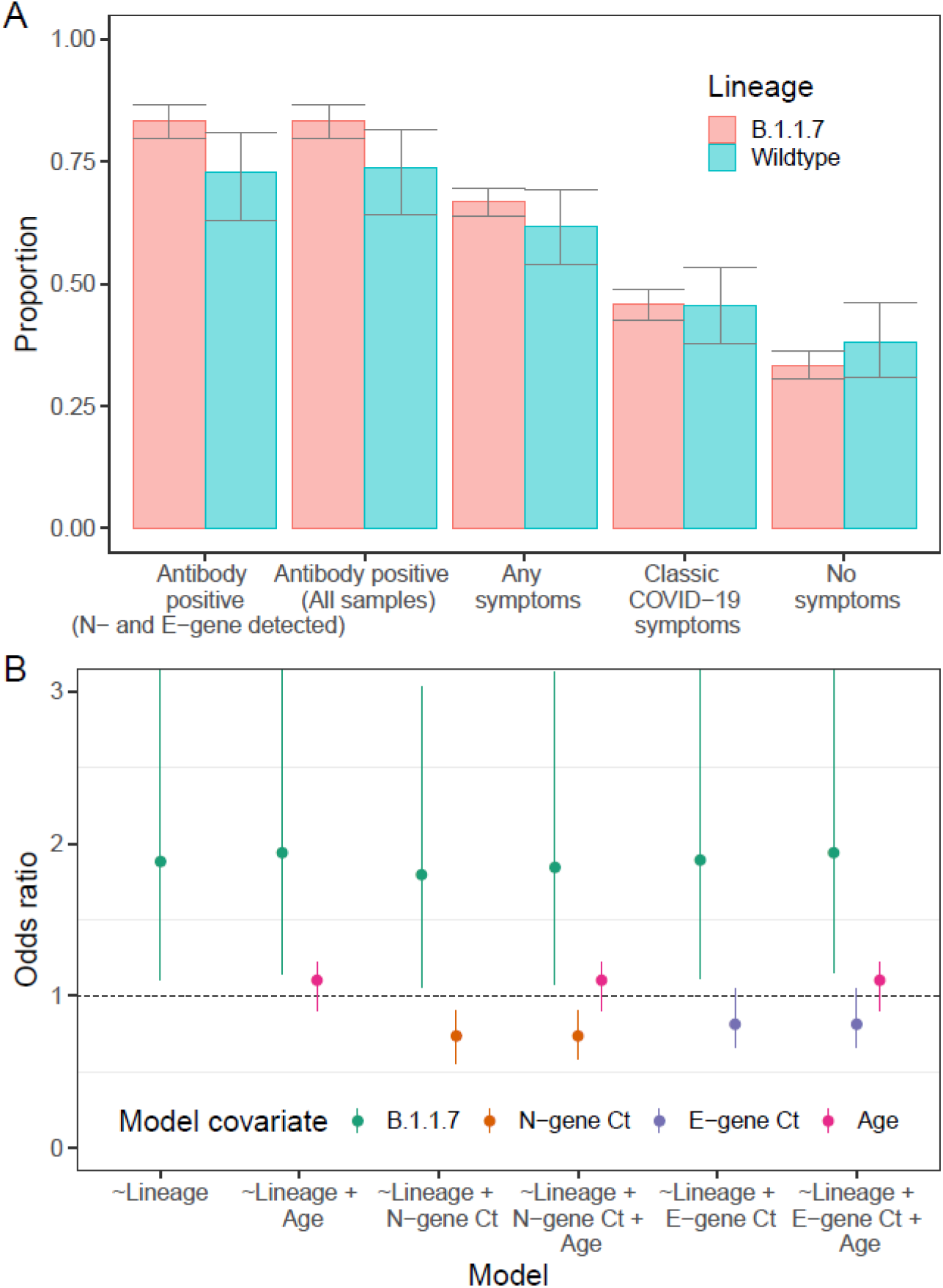
Symptoms and antibody positivity. (A) Proportion of those infected testing positive for antibodies 6 weeks after swab test (for all samples and for those that had both N- and E-gene detected), displaying any symptoms in the week prior to their swab test, displaying classic COVID-19 symptoms (loss of sense of taste, loss of sense of smell, new persistent cough, fever) in the week prior to their test and displaying no symptoms. (B) Odds ratios of the covariates of multiple logistic regression models. Each model had the result of the LFIA antibody test as the outcome variable with different combinations of lineage, N-gene Ct, E-gene Ct and Age as the covariates. OR displayed for B.1.1.7 is relative to wildtype. OR displayed for N- and E-gene Ct is relative to a change in Ct of +5. OR displayed for age is relative to a change of +10 years in age.

### Differences in cycle threshold values

Quantitative PCR Ct values for N and E gene targets were lower for the B.1.1.7 lineage relative to the wildtype (Table 1, Supplementary Figure 2). Mean N gene Ct value was 1.33 (0.60, 2.06) lower *(P*<0.001) and mean E gene Ct value was 0.90 (0.14, 1.67) lower (*P*=0.020). These values are indicative of a higher viral load in those infected with B.1.1.7 with a decrease in Ct of 1 corresponding to an approximate twofold increase in viral load [33]. This result matches earlier work that suggested the B.1.1.7 lineage has higher viral loads than other circulating lineages [34]. However, it is possible that both analyses have been influenced by the high growth rate of B.1.1.7 during its emergence relative to other lineages which could lead to a difference in the observed distribution generated by differences in the average time since infection [35]. Also, given that our sample workflow ensures that lower Ct values are more likely to receive a lineage designation, a lower intrinsic Ct value for B.1.1.7 could have led to an overestimated proportion of B.1.1.7 in the community.

**Table 1.**
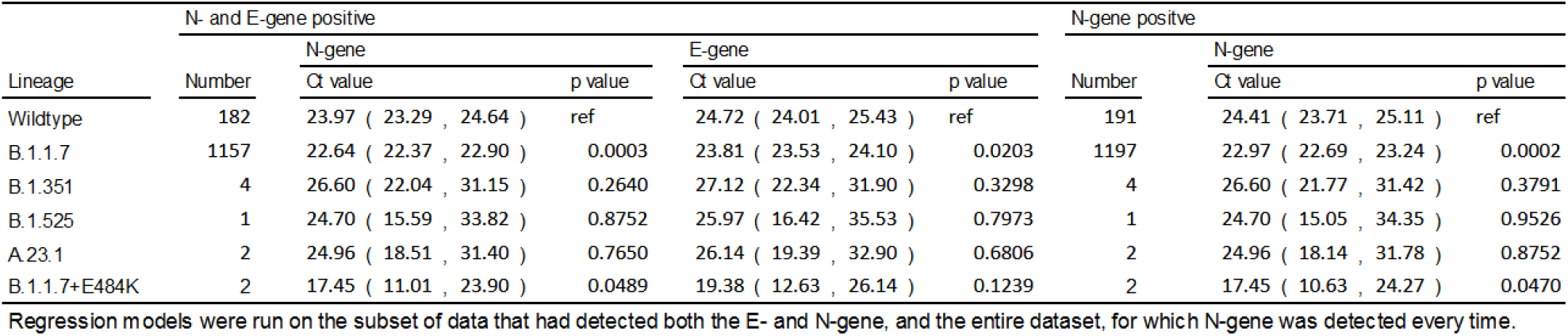
Results of Gaussian regression with either E-gene or N-gene Ct value as the observation and lineage as the explanatory variable.

### Differences in antibody positivity

Antibody positivity six weeks after the initial swab test, assessed using a lateral flow immunoassay [36], was higher in those infected with B.1.1.7 lineage relative to those infected with the wildtype lineage (Figure 3, Supplementary Table 4). Antibody positivity was 83.4% (79.7%, 86.5%) in those previously infected with B.1.1.7 and 72.8% (63.0%, 80.9%) in those previously infected with wildtype (*P*=0.018). This difference was not explained by patterns in N-gene Ct value, E-gene Ct value, or age (Figure 3). For example, the odds of B.1.1.7-positive participants being sero-positive were 1.84 (1.08, 3.13) higher than those who were wildtype-positive, using multivariable logistic regression that included N-gene Ct value and age as covariates. (Model 6, Supplementary Table 5). These differences in the likelihood of testing positive for antibodies may be caused by the immune response itself or by other unmeasured confounders such as the time from infection to swabbing.

### Variants of concern and variants under investigation

Though only a small number of B.1.351, B.1.525, A.23.1 and B.1.1.7+E484K were detected in January, because of the random sampling strategy used for REACT-1, we can estimate their prevalence with well-quantified uncertainty (Supplementary Table 6). The single detection of any lineage in round 8 corresponded to an estimated 812 (136, 4847) swab positive infections in England at any one time (Supplementary Table 6), suggesting that these lineages were already established in the community during January 2021. Additionally, none of the individuals infected with these lineages who answered the question reported that they had been abroad in the previous two weeks (Supplementary Table 7). However, not all of the participants who tested positive for a VOC or VUI answered the question about recent travel (one did not for B.1.525, one did not for A.23.1). Also, the sequences of some of the REACT-1 samples grouped very closely with other English isolates when compared to a representative global subsample of the lineage (e.g. ARCH-000045H2 for B.1.351 in Figure 4 and all isolates in Figure 5), further suggesting that significant local transmission was occurring at that time.

**Figure 4.**
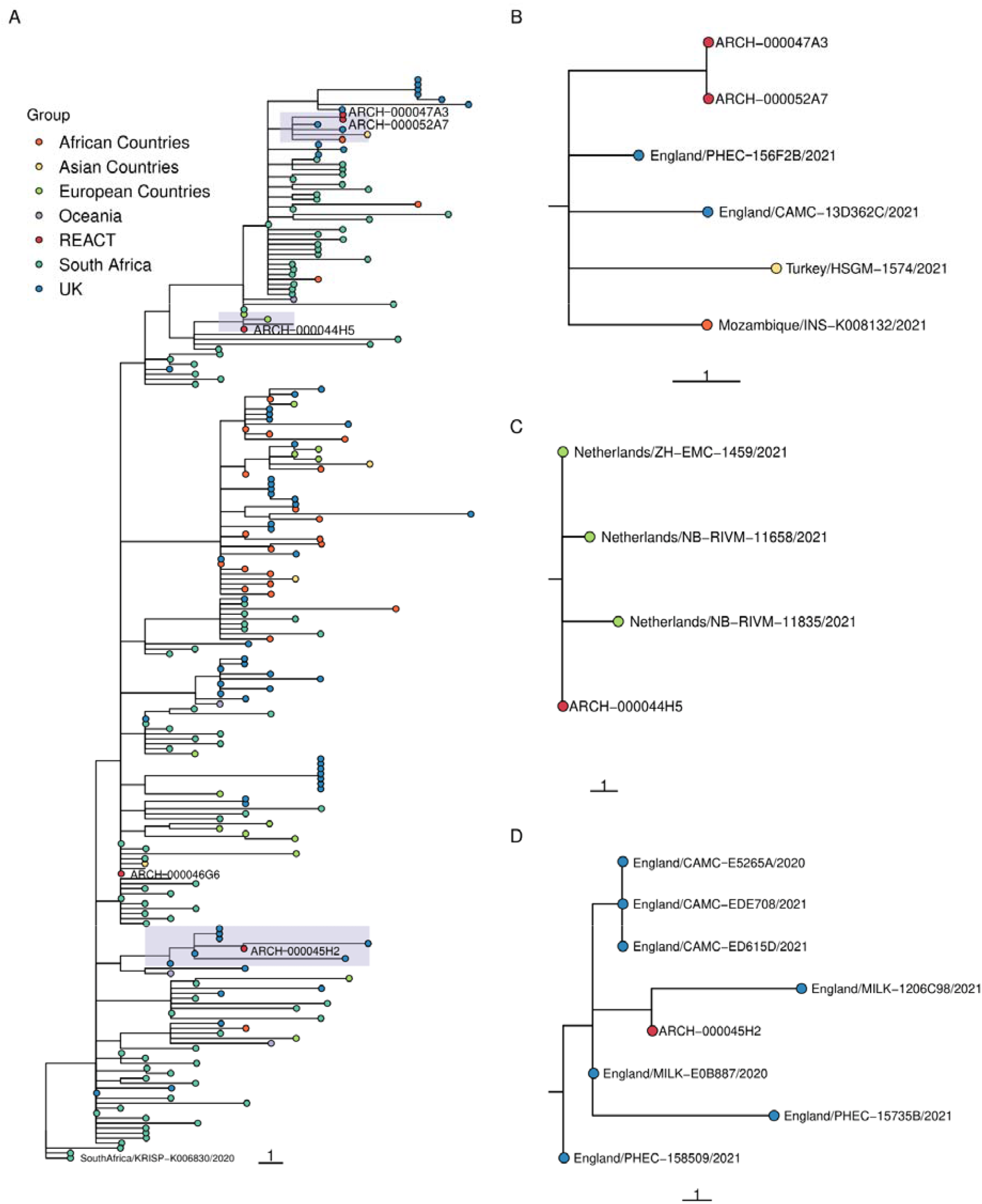
Phylogenetic tree showing relation of B.1.351 lineages detected in REACT-1 to other B.1.351 sequences in the COG-UK database. Sequences are coloured by the location in which the sequence was isolated. REACT lineages are coloured red and have an ID beginning with the sequence “ARCH-”-next to them. (A) Shows the subgroup of the entire constructed tree that contains all REACT sequences, re-rooted to the COG-UK sequence SouthAfrica/KRISP-K006830/2020. (B-D) show a zoomed-in view of the subtrees shown by the 3 shaded regions. Note that adjacent sequences ARCH-000047A3 and ARCH-000052A7 are multiple readings from the same individual.

**Figure 5.**
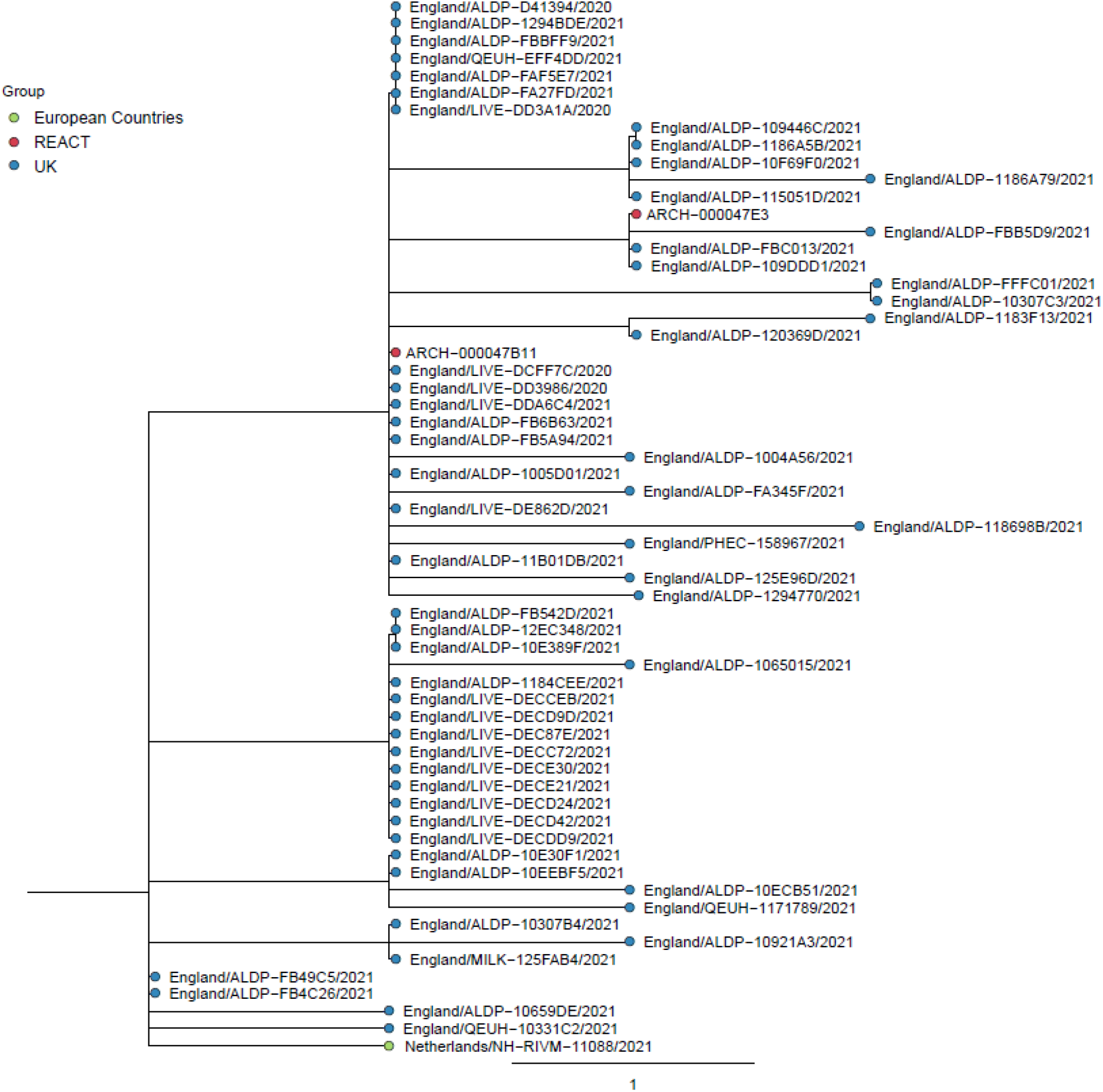
Phylogenetic tree showing relation of A.23.1 lineages detected in REACT-1 to other A.23.1 sequences in the COG-UK database. REACT-1 sequences are coloured in red and have an ID beginning with the sequence “ARCH-” next to them. All other sequences are coloured by the location in which the sequence was isolated.

Geographically, both samples of A.23.1 were detected in the North West (Figure 2, Supplementary Table 2). Given this lineage was first detected in England in the North West [16], these positive samples from community surveillance suggest that the lineage continued to circulate locally with limited spread to other regions of England. In contrast, B.1.1.7+E484K was detected in London and the North West, despite originally being detected in the South West [16], indicating either transmission out of the South West or that the E484K mutation arose independently within the B.1.1.7 lineage. The four B.1.351 samples were detected in London (1), South East (1), and East of England (2), and the single B.1.525 was also detected in London (Figure 2), suggesting that the capital and surrounding region plays an important role in the importation of lineages (see below Self-reported history of recent travel).

None of these VOCs or VOIs were detected in rounds 9 and 10 suggesting a decrease in their relative proportions. Fitting a logistic regression model to whether a sample was a specific VOC, VUI, or B.1.1.7, there was no evidence for a difference in transmissibility between A.23.1, B.1.525, B.1.1.7+E484K, and B.1.1.7. However, for B.1.351, which had the most samples available (n=4), the growth rate was estimated to be 0.110 (0.339, 0.002) less than B.1.1.7 (*P*=0.02) (Supplementary Table 8). The converse is seen in publicly available sequence data form community testing [37] (Supplementary Figure 3), which, for the same period of time, found that the frequency of B.1.351 grew faster than B.1.1.7 (Supplementary Table 8, Supplementary Figure 4). This difference likely reflects biases in public data, such as increased testing of international travellers and surge testing [38] in areas where variants are detected. The relative lineage dynamics do not seem to be consistent across space and time. Compared to patterns in England, higher proportions of B.1.351 relative to B.1.1.7 have been seen in some regions of Europe [12,14,39] and in Africa [7].

### Self-reported history of recent travel

Spatial patterns of observed VOCs and VOIs may be driven partly by geographical variation in the frequency with which people travel abroad. The overall proportion of participants reporting travel abroad in the past two weeks (Figure 6, Supplementary Table 9) in round 5 (September) was 1.63% (1.56%, 1.69%), but declined to 0.49% (0.46%, 0.53%) in round 8 (January), 0.11% (0.09%, 0.13%) in round 9 (February), and 0.10% (0.08%, 0.12%) in round 10 (March). London had the highest proportion and the South East the second highest for all rounds (Figure 6). We estimated that over 55% of the people returning from abroad to England during rounds 8, 9 and 10 were in London and the South East (Supplementary Table 9). Sub-regionally (Figure 6 and Supplementary Figure 5) we see that during round 5 (September) there was little spatial heterogeneity in the proportion of people who had been abroad two weeks prior, with similar proportions all across England. In contrast, during rounds 8, 9 and 10 there were high levels of heterogeneity with relatively higher proportions of travel among those living in central London and areas of Kent.

**Figure 6.**
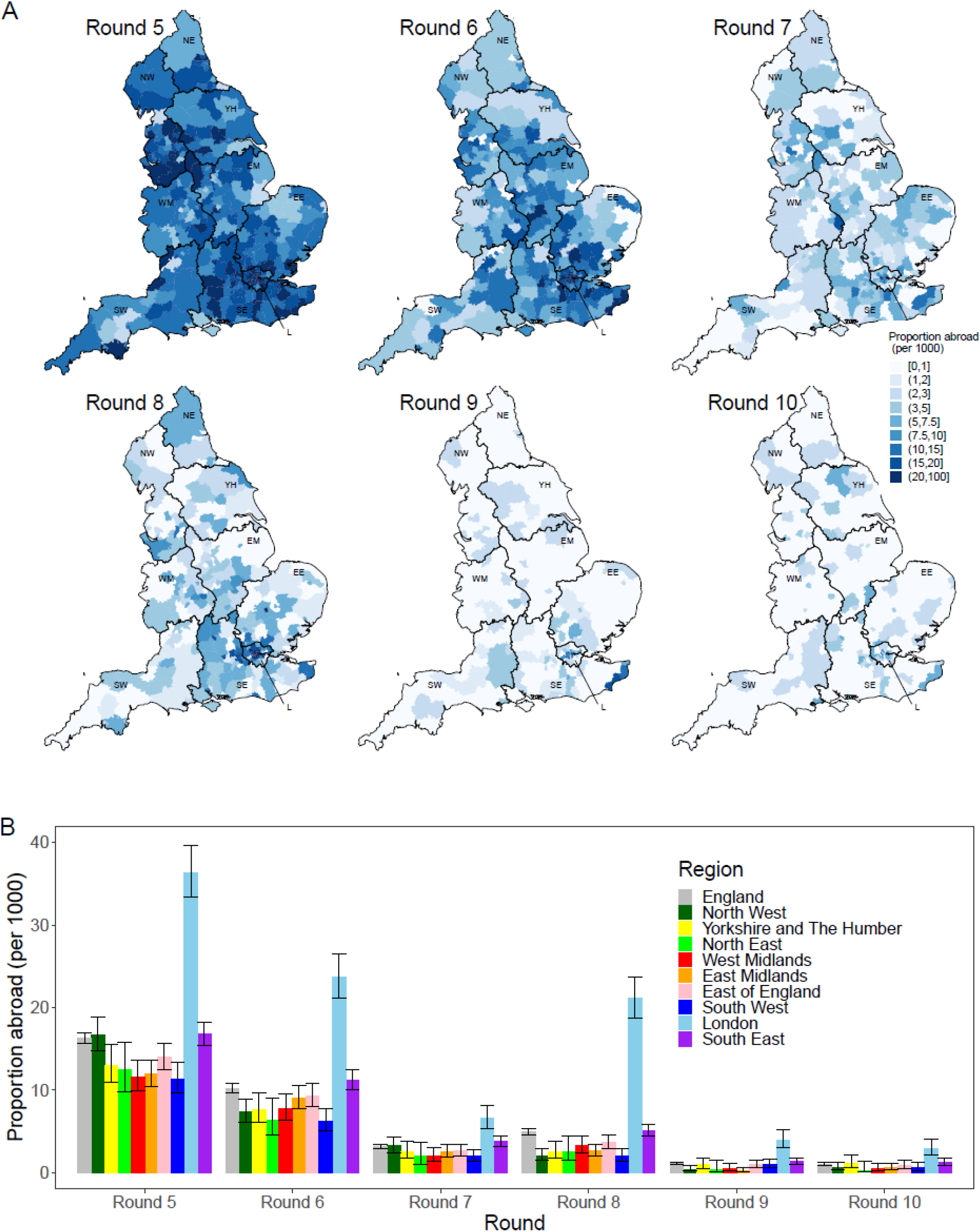
Patterns of frequency of returning from abroad in the prior two weeks. (A) Proportion of participants who answered they had been abroad in the previous two weeks by lower tier local authority. Regions: NE =North East, NW = North West, YH =Yorkshire and The Humber, EM = East Midlands, WM =West Midlands, EE = East ofEngland, L = London, SE = South East, SW = South West. (B) Proportion of individuals who answered that they had been abroad in the previous two weeks by region and round. Dates: Round 5 =18 September - 5 October 2020, Round 6 =16 October - 2 November 2020, Round 7 = 13 November - 3 December 2020, Round 8 = 6 January - 22 January 2021, Round 9 = 4 February - 23 February 2021, Round 10 = 11 March - 30 March 2021.

## Conclusion

We describe lineage dynamics for SARS-CoV-2 in England for the period January to March 2021, based on representative community samples. From January to March 2021, B.1.1.7 lineage continued to dominate the pandemic in England, further increasing in proportion over this time, spreading rapidly northwards and westwards. This may be explained by increased transmissibility over previously circulating lineages accompanied by a higher viral load.

Despite an increase in mortality observed for B.1.1.7 in other studies we find no evidence of a difference in the rate that infected individuals report the four classic COVID-19 symptoms (one or more of loss or change of sense of taste, loss or change of sense of smell, new persistent cough, fever) between B.1.1.7 and other circulating lineages.

Our results suggest other lineages were circulating in the community at lower levels at the beginning of the study period which coincided with the start of lockdown 3 in England. These were then out-competed by B.1.1.7 during the course of the study. In January, small numbers of B.1.351, B.1.525, A.23.1 and B.1.1.7+E484K were detected; although the numbers were small, due to the size and sampling method of the study, they likely indicate a substantial level of community transmission. In the later rounds in February and March none of these lineages were detected — the decline in proportion of B.1.351 relative to B.1.1.7 indicates that B.1.351 was, on average, less transmissible than B.1.1.7 over this period. The difference may have been partly a result of targeted public health interventions and reductions in foreign travel.

Our study has limitations. During round 8 a subsample of participants testing positive also undertook two additional swab tests. The sequencing results from these additional swabs indicates possible misallocation of samples to participants. Though not all of these samples contained sufficient viral copies for successful sequencing (high Ct) or not enough physical volume in the sample, we were able to sequence multiple swab tests for some participants. These extra sequences, when a lineage was determined, allowed augmentation of the data for round 8 for some of the participants whose first test was unable to be sequenced or have a lineage designated. Thirteen of the 175 participants who had multiple tests had discordant lineage designations (Supplementary Table 10). Four of these divergent lineage designations were not incongruent, for example B.1.351 and B.1, likely reflecting a lower quality second sequence. For these four cases, the more advanced lineage was selected (B.1.351 over B.1). The remaining nine lineages could not be determined definitively, and so have been removed from the main analysis. This points to some potential sample mix-ups caused by manual cherry picking in the diagnostic pipeline; however, these errors are only likely to affect the most prevalent lineages, specifically B.1.1.7 and B.1.177.

The success of B.1.1.7 against the prior wild type and more recent variants during the first three months of 2021 in England during lockdown does not necessarily mean that future imported variants such as B.1.617.2 (first identified in India) will not succeed [40]. The immune landscape [41] against SARS-CoV-2 in the UK is changing rapidly because of high uptake of effective vaccines across a broad range of age groups. On the other hand, as stringent interventions during lockdown are eased, the potential for person-to-person contact and overall average transmissibility of SARS-CoV-2 in England will increase.

## Methods

Methods for the REACT-1 study have been described previously [27]. Since May 2020, there have been 10 rounds of data collection approximately every month with between 140,000 and 175,000 swab tests completed over a 2-3 week period by a random subset of the population of England aged 5 and over. From round 8 onwards, all positive tests with a low N-gene Ct value (less than 34 used initially, but criteria changed midway through round 9 to less than 32 due to high rate of sequencing failure in those with N-gene Ct>32 [approx 88%]) and a high enough volume were sent for genome sequencing (Public Health England Research Ethics Governance Group (reference: R&D NR0195)). Extracted RNA was amplified using the ARTIC protocol [43] with sequencing libraries prepared using CoronaHiT [44], and sequenced on the Illumina NextSeq 500 platform. Each set of 96 samples included one positive and one negative control. Raw sequencing data were analysed using the ARTIC bioinformatic pipeline [45] and uploaded to CLIMB [46] for further analysis.

From the genome sequences lineages are assigned using a machine learning based assignment algorithm, PangoLEARN [47] (database version 2021-04-28) with lineage assignment following the Pangolin nomenclature [9]. Not all obtained sequences were of a high enough quality for a lineage to be determined and so are not included in the analysis. Further, samples in which less than 50% of bases were covered were excluded from the analysis. A diagram showing how many positive samples were sequenced and how many had a lineage determined is shown in Supplementary Figure 1.

Following lineage allocation by the algorithm, each sequence was then also investigated individually, particularly for the presence of lineage defining SNPs. This allowed for lineages that were too low quality to call by PangoLEARN to be manually assigned. This occurred twice, once for a B.1 lineage call that exhibited six of 14 B.1.351 lineage defining mutations (adjusted to B.1.351), and once for a B.1 lineage call that exhibited 11 out of 16 B.1.525 lineage defining mutations (adjusted to B.1.525). In depth analysis of low coverage regions of both samples which fell below the normal minimum threshold of 10X coverage showed that all lineage defining mutations were present in at least one sequence read, further supporting these probable lineage designations. The thresholds for ‘probable’ lineage designations are defined by Public Health England [48]. B.1.1.7 lineages that also had the E484K SNP present were designated to a separate lineage (B.1.1.7+E484K).

Phylogenetic trees were constructed in order to investigate how detected VOCs and VUIs fit into the wider epidemic context. A subsample of sequences for each variant was selected from a curated database of COG-UK up to 12 April 2021. A representative subsample for each lineage was selected using Civet [49] with a collapse threshold of 2. Two hundred and twelve of 1583 B.1.351 sequences and 21 of 60 A.23.1 sequences were selected. For each variant the maximum-likelihood phylogenetic tree was constructed using a HKY model implemented using IQ-TREE [50].

95% confidence intervals in lineage proportions were calculated assuming a binomial distribution using the Wilson method [51], which is preferred when the number of positive outcomes is small [52]. Differences in multinomial proportions between rounds were assessed using a multinomial goodness-of-fit test implemented using the XNomial package in R [53].

Potential confounding effects of region and age group on B.1.1.7 proportion during round 8 were investigated using logistic regression with a binomial likelihood and logit link function. Lineage assignment of B.1.1.7 versus any other was the binary outcome variable and both region and age group were included as covariates. Similar analysis was not attempted on round 9 and round 10 due to the small number of non-B.1.1.7 lineages.

Estimates of the average true number of swab positive cases by lineage at any one time during rounds 8, 9 and 10 were calculated by multiplying the estimates of weighted prevalence for rounds 8, 9 and 10 [54], the proportion of each lineage for rounds 8, 9 and 10 (Supplementary Table 1 and Supplementary Table 2), and the population size of England and each region [55]

Relative differences in growth rates between two lineages were estimated by fitting a Bayesian logistic regression model to the binary lineage outcome. This was converted into an additive difference (□) in R through the equation □□ = □□ × <u>□</u>, with the assumption □ = 1 + □ × □ [56] where r is the growth rate of a lineage and □ is the mean generation time, assumed to be 6.29 days [57] for both lineages.

Smoothed spatial estimates of the relative proportion of two lineages were estimated using a Bayesian generalised-linear mixed-effects model implemented in the R package glmmfields [58]. We included 25 knots to describe the spatial processes and random spatial effects were assumed to follow a multivariate-t distribution. Priors of the model were chosen to be uninformative.

A subsample of positive participants in round 8 underwent a lateral flow immunoassay [36] approximately 6 weeks after their initial swab test. Differences in raw antibody positivity by lineages were assessed using logistic regression, with a binomial likelihood using a logit link function, and the result of the antibody test (positive/negative) as the binary outcome variable. Regression was performed using the subset of the data in which both the N-gene and E-gene had been detected. Further regression models were performed including different combinations of age, N-gene Ct value and E-gene Ct value as additional covariates. Further exploratory analyses were conducted with models including interaction terms between different combinations of variables and smoothed terms for some variables (not reported).

## Supporting information

COG-UK Author List

## Data Availability

Assembled/consensus genomes are available from GISAID subject to minimum quality control criteria. Raw reads are available from European Nucleotide Archive (ENA). All genomes, phylogenetic trees, and basic metadata are available from the COG-UK consortium website (https://www.cogconsortium.uk). For confidentiality reasons, extended metadata are not publicly available, however some may be available upon request. Supplementary tables are available in the spreadsheet (link provided below)

https://docs.google.com/spreadsheets/d/1Fk-IDQFc6eG4lV2j8_dnYHk8zuG3Fb-Pfwitnxx82mk/edit?usp=sharing

## Acknowledgements

SR, CAD acknowledge support: Medical Research Council (MRC) Centre for Global Infectious Disease Analysis, National Institute for Health Research (NIHR) Health Protection Research Unit (HPRU), Wellcome Trust (200861/Z/16/Z, 200187/Z/15/Z), and Centres for Disease Control and Prevention (US, U01CK0005-01-02). GC is supported by an NIHR Professorship. HW acknowledges support from an NIHR Senior Investigator Award and the Wellcome Trust (205456/Z/16/Z). PE is Director of the MRC Centre for Environment and Health (MR/L01341X/1, MR/S019669/1). PE acknowledges support from Health Data Research UK (HDR UK); the NIHR Imperial Biomedical Research Centre; NIHR HPRUs in Chemical and Radiation Threats and Hazards, and Environmental Exposures and Health; the British Heart Foundation Centre for Research Excellence at Imperial College London (RE/18/4/34215); and the UK Dementia Research Institute at Imperial (MC_PC_17114). We thank The Huo Family Foundation for their support of our work on COVID-19. AJP, JOG gratefully acknowledge the support of the Biotechnology and Biological Sciences Research Council (BBSRC); their research was funded by the BBSRC Institute Strategic Programme Microbes in the Food Chain BB/R012504/1 and its constituent project BBS/E/F/000PR10352. We thank members of the COVID-19 Genomics UK (COG-UK) Consortium UK for their contributions to generating the genomic data used in this study. COG-UK is supported by funding from the MRC (part of UK Research & Innovation ([UKRI]), the NIHR and Genome Research Limited, operating as the Wellcome Sanger Institute.

We thank key collaborators on this work – Ipsos MORI: Kelly Beaver, Sam Clemens, GaryWelch, Nicholas Gilby, Kelly Ward and Kevin Pickering; Institute of Global Health Innovationat Imperial College: Gianluca Fontana, Didi Thompson and Lenny Naar; Molecular Diagnostic Unit, Imperial College London: Prof. Graham Taylor; Patient Experience Research Centre at Imperial College and the REACT Public Advisory Panel; NHS Digital for access to the NHS register; and the Department of Health and Social Care for logistic support.

## Ethics

The COG-UK study protocol was approved by the Public Health England Research Ethics Governance Group (reference: R&D NR0195). This study was conducted as part of surveillance for COVID-19 infections under the auspices of Section 251 of the NHS Act 2006 and/or Regulation 3 of The Health Service (Control of Patient Information) Regulations 2002. The REACT-1 study received research ethics approval from the South Central-Berkshire BResearch Ethics Committee (IRAS ID: 283787).

## Data availability

Assembled/consensus genomes are available from GISAID subject to minimum quality control criteria. Raw reads are available from European Nucleotide Archive (ENA). All genomes, phylogenetic trees, and basic metadata are available from the COG-UK consortium website (https://www.cogconsortium.uk). For confidentiality reasons, extended metadata are not publicly available, however some may be available upon request.

## Materials & Correspondence

Correspondence and requests for materials should be addressed to Steven Riley and Paul Elliott, School of Public Health, Imperial College London, Norfolk Place, London, W2 1PG

## Additional information

Titles for <u>Supplementary Tables 1 to 10</u> are included below with the tables themselves in this <u>spreadsheet</u>. <u>Supplementary Figures 1 to 5</u> are included below. Full list of <u>COG-UK author’s names</u> and affiliations are available in this <u>spreadsheet</u>.

## Supplementary Tables and Figures

**Supplementary Table 1**. Lineages detected in round 8, round 9 and round 10 of REACT-1.

Supplementary Table 1 is available in this <u>spreadsheet</u>

**Supplementary Table 2**. Regional distribution of positives for lineages in England for round 8, 9 and 10.

Supplementary Table 2 is available in this <u>spreadsheet</u>

**Supplementary Table 3**. Symptom status by lineage

Supplementary Table 3 is available in this <u>spreadsheet</u>

**Supplementary Table 4**. Antibody positivity 6 weeks after a positive swab test by lineage type.

Supplementary Table 4 is available in this <u>spreadsheet</u>

**Supplementary Table 5**. Multivariate logistic regression models to determine the effect of lineage on antibody positivity.

Supplementary Table 5 is available in this <u>spreadsheet</u>

**Supplementary Table 6**. Estimates of the average true number of swab positive cases by lineage at any one time during round 8, 9 and 10 in England and in each region of England.

Supplementary Table 6 is available in this <u>spreadsheet</u>

**Supplementary Table 7**. Number of participants self-reporting being abroad in the two weeks prior to taking their swab test by round and lineage type.

Supplementary Table 7 is available in this <u>spreadsheet</u>

**Supplementary Table 8**. Implied difference in growth rates of detected VOCs and VUIs. Supplementary Table 8 is available in this <u>spreadsheet</u>

**Supplementary Table 9**. Proportion of participants that reported being abroad two weeks prior to their swab test by region and by round, estimated average number of people who had been abroad in the previous two weeks by region and round, and the overall percentage of people returning from abroad that were in each region of England.

Supplementary Table 9 is available in this <u>spreadsheet</u>

**Supplementary Table 10**. Description of the participants who had discordant lineage designations and the overall lineage designation made.

Supplementary Table 10 is available in this <u>spreadsheet</u>

**Supplementary Figure 1.**
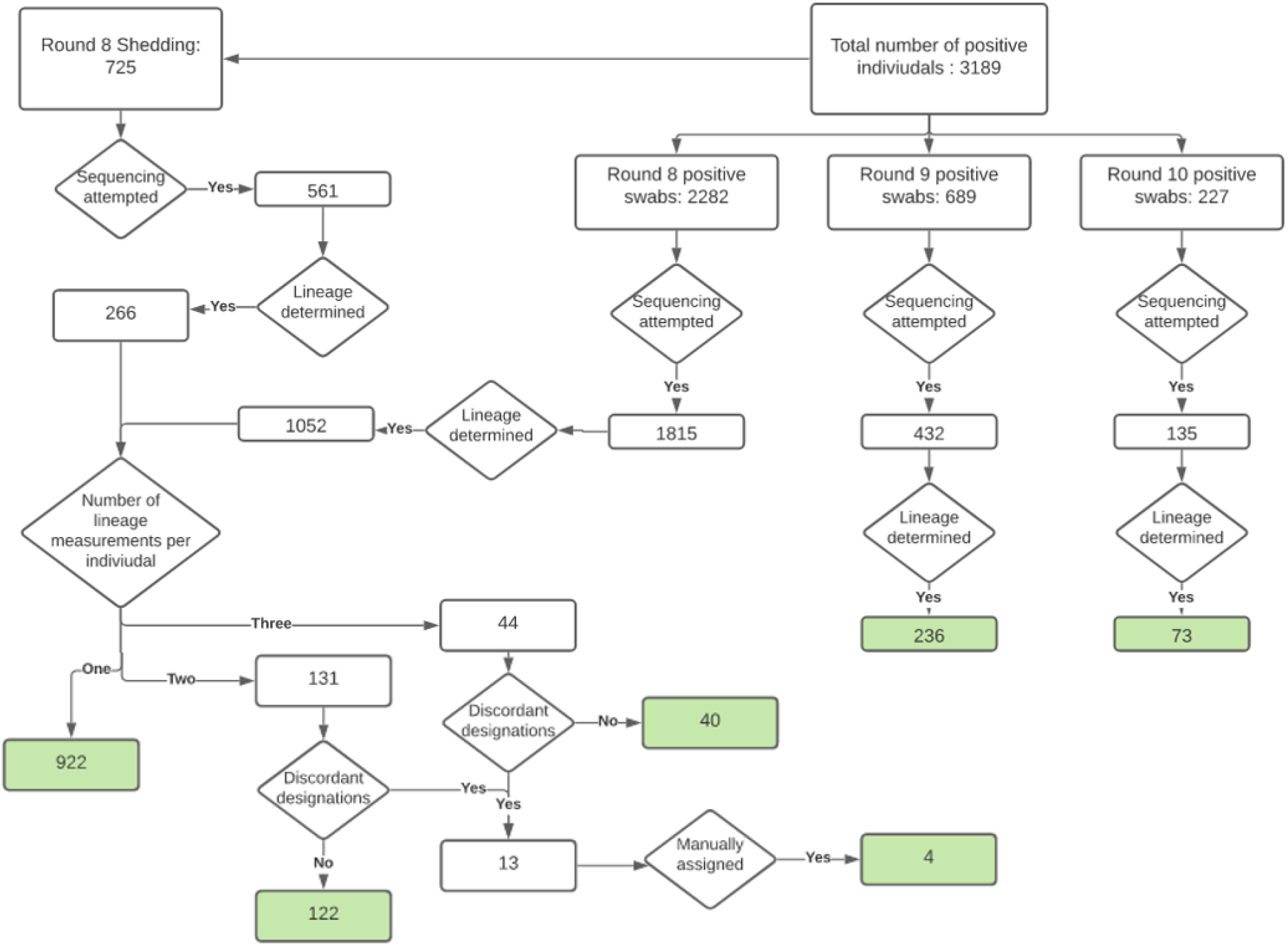
Diagram showing the number of positive samples, the number for which sequencing was attempted, and the number of lineages that were determined and therefore used in the analysis (Green) for rounds 8, 9 and 10 of the study.

**Supplementary Figure 2.**
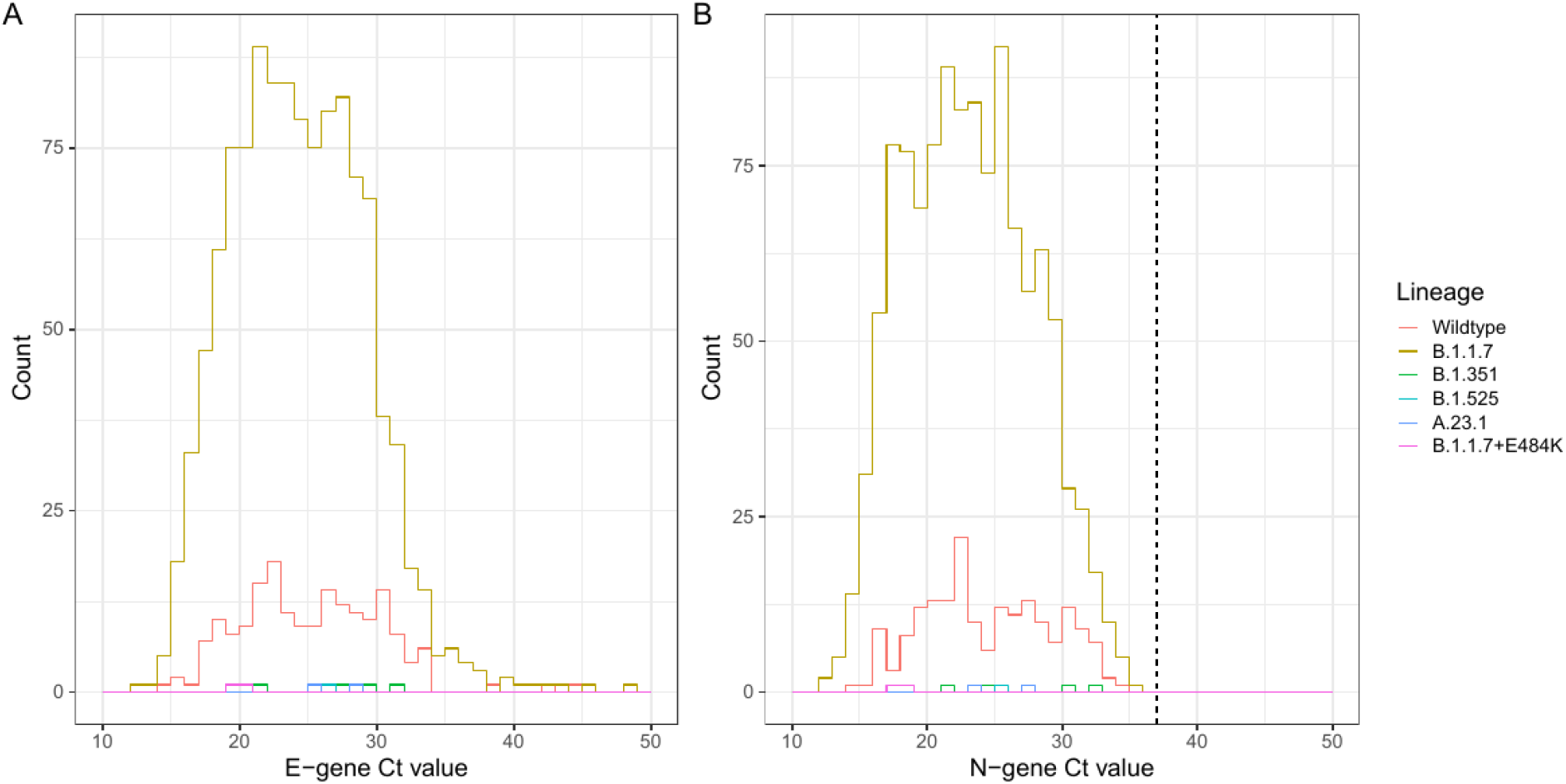
Distribution of Ct values by lineage for (A) E-gene and (B) N-gene. Dotted line shows the N-gene Ct cutoff value of 37 that was used to define positivity.

**Supplementary Figure 3.**
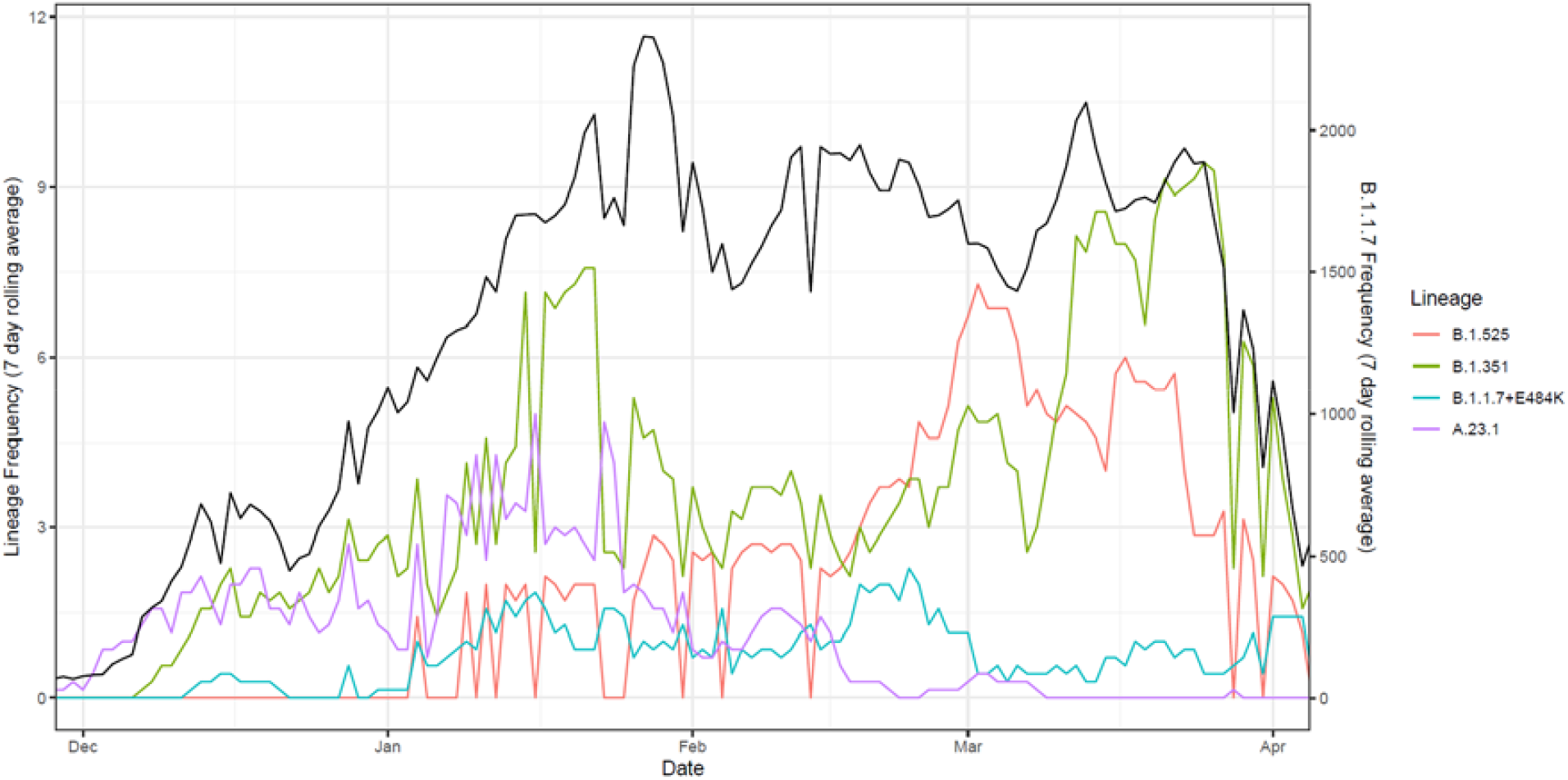
Central 7 day rolling average of the frequency of B.1.1.7 (Black, right y-axis), B.1.351 (Green, left y-axis), B.1.525 (Red, left y-axis), A.23.1 (Purple, left y-axis) and B.1.1.7+E484K (Blue, left y-axis) in publicly available data.

**Supplementary Figure 4.**
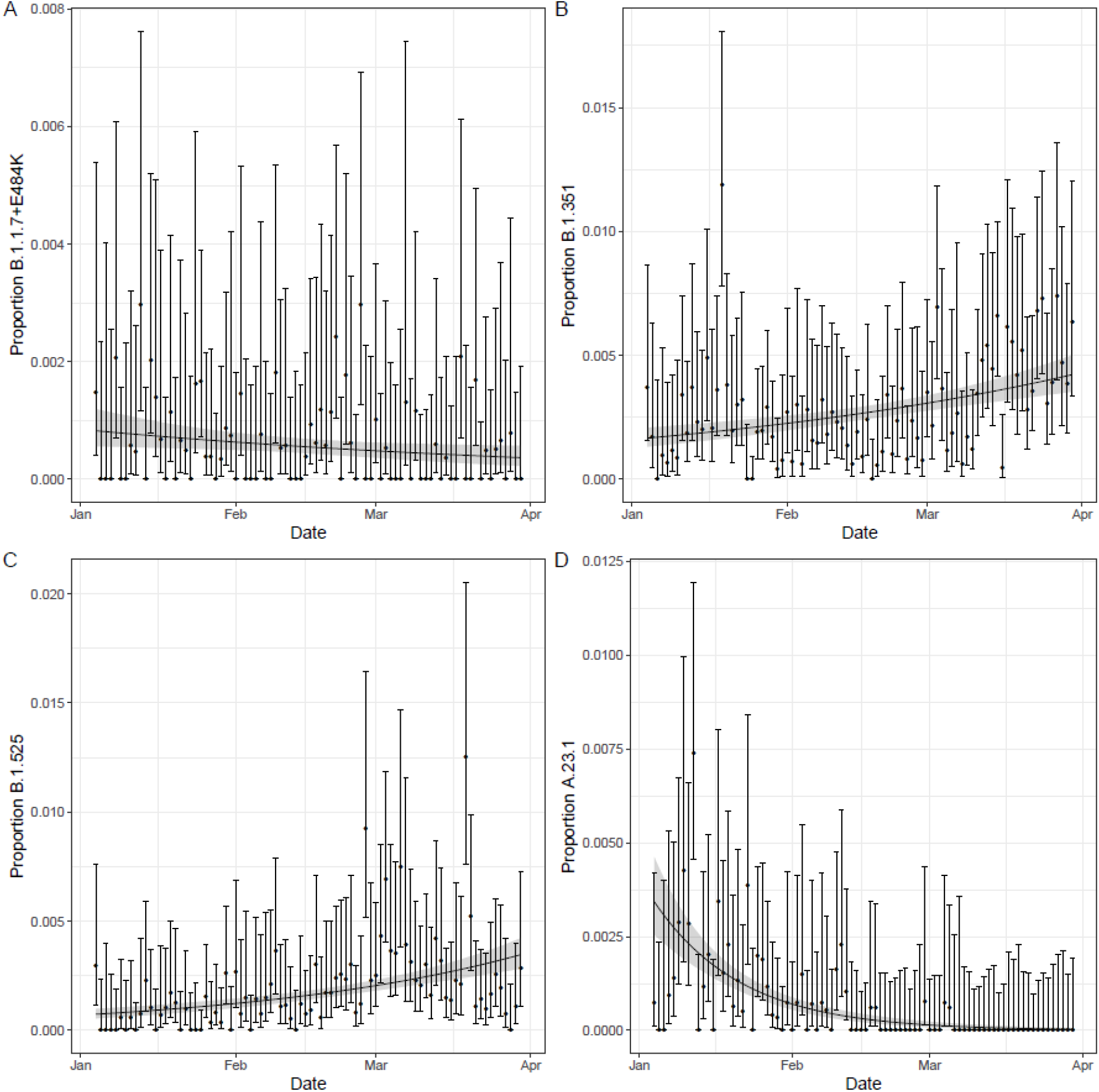
Plot of the proportion of (A) B.1.1.7+E484K (B) B.1.351 (C) B.1.525 and (D) A.23.1 relative to B.1.1.7 lineage in publicly available data over the same time span as the REACT-1 rounds 8 to 10 lineage data. Error bars show the 95% confidence interval for each daily proportion calculation. Shaded region shows best fit Bayesian logistic regression model with 95% credible interval.

**Supplementary Figure 5.**
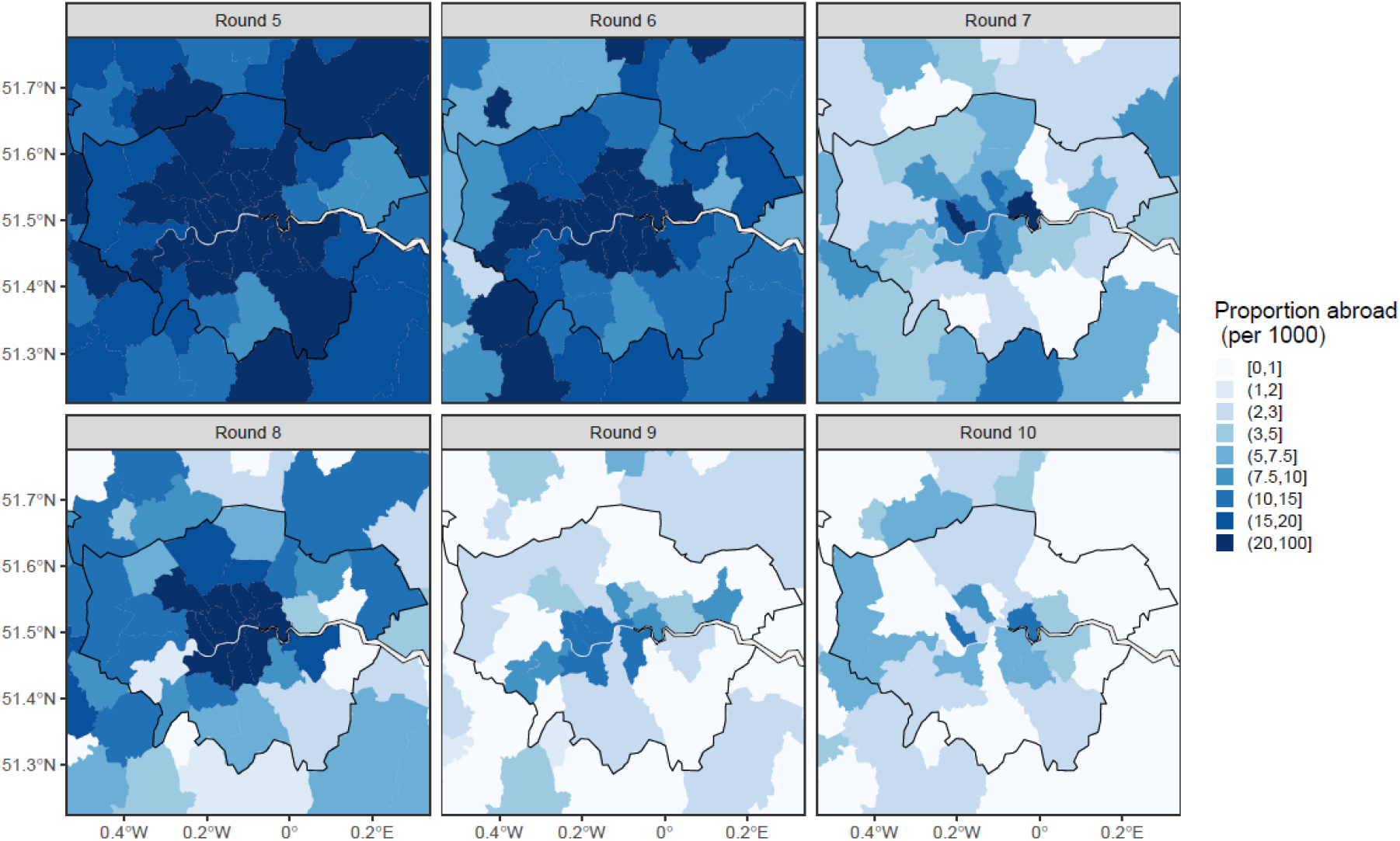
Proportion of participants who answered they had been abroad in the previous two weeks by lower tier local authority zoomed into the London region. Dates: Round 5 =18 September - 5 October 2020, Round 6 =16 October - 2 November 2020, Round 7 = 13 November - 3 December 2020, Round 8 = 6 January - 22 January 2021, Round 9 = 4 February - 23 February 2021, Round 10 = 11 March - 30 March 2021.

