## Supplementary material for "SARS-CoV-2 lineage dynamics in England from January to March 2021 inferred from representative community samples": COG-UK Author List

**Funding acquisition,** **Leadership and supervision,** **Metadata curation,** **Project administration,** **Samples and logistics,** **Sequencing and analysis,** **Software and analysis tools, and Visualisation:**

Dr Samuel C Robson ^13^.

**Funding acquisition,** **Leadership and supervision,** **Metadata curation,** **Project administration,** **Samples and logistics,** **Sequencing and analysis, and Software and analysis tools:**

Prof Nicholas J Loman ^41^, Dr Thomas R Connor ^10,^ ^69^.

**Leadership and supervision,** **Metadata curation,** **Project administration,** **Samples and logistics,** **Sequencing and analysis,** **Software and analysis tools, and Visualisation:**

Dr Tanya Golubchik ^5^.

**Funding acquisition,** **Metadata curation,** **Samples and logistics,** **Sequencing and analysis,** **Software and analysis tools, and Visualisation:**

Dr Rocio T Martinez Nunez ^42^.

**Funding acquisition,** **Leadership and supervision,** **Metadata curation,** **Project administration, and Samples and logistics:**

Dr Catherine Ludden ^88^.

**Funding acquisition,** **Leadership and supervision,** **Metadata curation,** **Samples and logistics, and Sequencing and analysis:**

Dr Sally Corden ^69^.

**Funding acquisition,** **Leadership and supervision,** **Project administration,** **Samples and logistics, and Sequencing and analysis:**

Ian Johnston ^99^ and Dr David Bonsall ^5^.

**Funding acquisition,** **Leadership and supervision,** **Sequencing and analysis,** **Software and analysis tools, and Visualisation:**

Prof Colin P Smith ^87^ and Dr Ali R Awan ^28^.

**Funding acquisition,** **Samples and logistics,** **Sequencing and analysis,** **Software and analysis tools, and Visualisation:**

Dr Giselda Bucca ^87^.

**Leadership and supervision,** **Metadata curation,** **Project administration,** **Samples and logistics, and Sequencing and analysis:**

Dr M. Estee Torok ^22,^ ^101^.

**Leadership and supervision,** **Metadata curation,** **Project administration,** **Samples and logistics, and Visualisation:**

Dr Kordo Saeed ^81,^ ^110^ and Dr Jacqui A Prieto ^83,^ ^109^.

**Leadership and supervision,** **Metadata curation,** **Project administration,** **Sequencing and analysis, and Software and analysis tools:**

Dr David K Jackson ^99^.

**Metadata curation,** **Project administration,** **Samples and logistics,** **Sequencing and analysis, and Software and analysis tools:**

Dr William L Hamilton ^22^.

**Metadata curation,** **Project administration,** **Samples and logistics,** **Sequencing and analysis, and Visualisation:**

Dr Luke B Snell ^11^.

**Funding acquisition,** **Leadership and supervision,** **Metadata curation, and Samples and logistics:**

Dr Catherine Moore ^69^.

**Funding acquisition,** **Leadership and supervision,** **Project administration,**and **Samples and logistics:**

Dr Ewan M Harrison ^99,^ ^88^.

**Leadership and supervision,** **Metadata curation,** **Project administration, and Samples and logistics:**

Dr Sonia Goncalves ^99^.

**Leadership and supervision,** **Metadata curation,** **Samples and logistics, and Sequencing and analysis:**

Prof Ian G Goodfellow ^24^, Dr Derek J Fairley ^3,^ ^72^, Prof Matthew W Loose ^18^ and Joanne Watkins ^69^.

**Leadership and supervision,** **Metadata curation,** **Samples and logistics, and Software and analysis tools:**

Rich Livett ^99^.

**Leadership and supervision,** **Metadata curation,** **Samples and logistics, and Visualisation:**

Dr Samuel Moses ^25,^ ^106^.

**Leadership and supervision,** **Metadata curation,** **Sequencing and analysis, and Software and analysis tools:**

Dr Roberto Amato ^99^, Dr Sam Nicholls ^41^ and Dr Matthew Bull ^69^.

**Leadership and supervision,** **Project administration,** **Samples and logistics, and Sequencing and analysis:**

Prof Darren L Smith ^37,^ ^58,^ ^105^.

**Leadership and supervision,** **Sequencing and analysis,** **Software and analysis tools, and Visualisation:**

Dr Jeff Barrett ^99^ and Prof David M Aanensen ^14,^ ^114^.

**Metadata curation,** **Project administration,** **Samples and logistics, and Sequencing and analysis:**

Dr Martin D Curran ^65^, Dr Surendra Parmar ^65^, Dr Dinesh Aggarwal ^95,^ ^99,^ ^64^ and Dr James G Shepherd ^48^.

**Metadata curation,** **Project administration,** **Sequencing and analysis, and Software and analysis tools:**

Dr Matthew D Parker ^93^.

**Metadata curation,** **Samples and logistics,** **Sequencing and analysis, and Visualisation:**

Dr Sharon Glaysher ^61^.

**Metadata curation,** **Sequencing and analysis,** **Software and analysis tools, and Visualisation:**

Dr Matthew Bashton ^37,^ ^58^, Dr Anthony P Underwood ^14,^ ^114^, Dr Nicole Pacchiarini ^69^ and Dr Katie F Loveson ^77^.

**Project administration,** **Sequencing and analysis,** **Software and analysis tools, and Visualisation:**

Dr Alessandro M Carabelli ^88^.

**Funding acquisition,** **Leadership and supervision, and Metadata curation:**

Dr Kate E Templeton ^53,^ ^90^.

**Funding acquisition,** **Leadership and supervision, and Project administration:**

Dr Cordelia F Langford ^99^, John Sillitoe ^99^, Dr Thushan I de Silva ^93^ and Dr Dennis Wang ^93^.

**Funding acquisition,** **Leadership and supervision, and Sequencing and analysis:**

Prof Dominic Kwiatkowski ^99,^ ^107^, Prof Andrew Rambaut ^90^, Dr Justin O’Grady ^70,^ ^89^ and Dr Simon Cottrell ^69^.

**Leadership and supervision,** **Metadata curation, and Sequencing and analysis:**

Prof Matthew T.G. Holden ^68^ and Prof Emma C Thomson ^48^.

**Leadership and supervision,** **Project administration, and Samples and logistics:**

Dr Husam Osman ^64,^ ^36^, Dr Monique Andersson ^59^, Prof Anoop J Chauhan ^61^ and Dr Mohammed O Hassan-Ibrahim ^6^.

**Leadership and supervision,** **Project administration, and Sequencing and analysis:**

Dr Mara Lawniczak ^99^.

**Leadership and supervision,** **Samples and logistics, and Sequencing and analysis:**

Prof Ravi Kumar Gupta ^88,^ ^113^, Dr Alex Alderton ^99^, Dr Meera Chand ^66^, Dr Chrystala Constantinidou ^94^, Dr Meera Unnikrishnan ^94^, Prof Alistair C Darby ^92^, Prof Julian A Hiscox ^92^ and Prof Steve Paterson ^92^.

**Leadership and supervision,** **Sequencing and analysis, and Software and analysis tools:**

Dr Inigo Martincorena ^99^, Prof David L Robertson ^48^, Dr Erik M Volz ^39^, Dr Andrew J Page ^70^ and Prof Oliver G Pybus ^23^.

**Leadership and supervision,** **Sequencing and analysis, and Visualisation:**

Dr Andrew R Bassett ^99^.

**Metadata curation,** **Project administration, and Samples and logistics:**

Dr Cristina V Ariani ^99^, Dr Michael H Spencer Chapman ^99,^ ^88^, Dr Kathy K Li ^48^, Dr Rajiv N Shah ^48^, Dr Natasha G Jesudason ^48^ and Dr Yusri Taha ^50^.

**Metadata curation,** **Project administration, and Sequencing and analysis:**

Martin P McHugh ^53^ and Dr Rebecca Dewar ^53^.

**Metadata curation,** **Samples and logistics, and Sequencing and analysis:**

Dr Aminu S Jahun ^24^, Dr Claire McMurray ^41^, Ms Sarojini Pandey ^84^, Dr James P McKenna ^3^, Dr Andrew Nelson ^58,^ ^105^, Dr Gregory R Young ^37,^ ^58^, Dr Clare M McCann ^58,^ ^105^ and Mr Scott Elliott ^61^.

**Metadata curation,** **Samples and logistics, and Visualisation:**

Ms Hannah Lowe ^25^.

**Metadata curation,** **Sequencing and analysis, and Software and analysis tools:**

Dr Ben Temperton ^91^, Dr Sunando Roy ^82^, Dr Anna Price ^10^, Dr Sara Rey ^69^ and Mr Matthew Wyles ^93^.

**Metadata curation,** **Sequencing and analysis, and Visualisation:**

Stefan Rooke ^90^ and Dr Sharif Shaaban ^68^.

**Project administration,** **Samples and logistics,** **Sequencing and analysis:**

Dr Mariateresa de Cesare ^98^.

**Project administration,** **Samples and logistics, and Software and analysis tools:**

Laura Letchford ^99^.

**Project administration,** **Samples and logistics, and Visualisation:**

Miss Siona Silveira ^81^, Dr Emanuela Pelosi ^81^ and Dr Eleri Wilson-Davies ^81^.

**Samples and logistics,** **Sequencing and analysis, and Software and analysis tools:**

Dr Myra Hosmillo ^24^.

**Sequencing and analysis,** **Software and analysis tools, and Visualisation:**

Áine O'Toole ^90^, Dr Andrew R Hesketh ^87^, Mr Richard Stark ^94^, Dr Louis du Plessis ^23^, Dr Chris Ruis ^88^, Dr Helen Adams ^4^ and Dr Yann Bourgeois ^76^.

**Funding acquisition, and Leadership and supervision:**

Dr Stephen L Michell ^91^, Prof Dimitris Grammatopoulos ^84,^ ^112^, Dr Jonathan Edgeworth ^12^, Prof Judith Breuer ^30,^ ^82^, Prof John A Todd ^98^ and Dr Christophe Fraser ^5^.

**Funding acquisition, and Project administration:**

Dr David Buck ^98^ and Michaela John ^9^.

**Leadership and supervision, and Metadata curation:**

Dr Gemma L Kay ^70^.

**Leadership and supervision, and Project administration:**

Steve Palmer ^99^, Prof Sharon J Peacock ^88,^ ^64^ and David Heyburn ^69^.

**Leadership and supervision, and Samples and logistics:**

Danni Weldon ^99^, Dr Esther Robinson ^64,^ ^36^, Prof Alan McNally ^41,^ ^86^, Dr Peter Muir ^64^, Dr Ian B Vipond ^64^, Dr John BoYes ^29^, Dr Venkat Sivaprakasam ^46^, Dr Tranprit Saluja ^75^, Dr Samir Dervisevic ^54^ and Dr Emma J Meader ^54^.

**Leadership and supervision, and Sequencing and analysis:**

Dr Naomi R Park ^99^, Karen Oliver ^99^, Dr Aaron R Jeffries ^91^, Dr Sascha Ott ^94^, Dr Ana da Silva Filipe ^48^, Dr David A Simpson ^72^ and Dr Chris Williams ^69^.

**Leadership and supervision, and Visualisation:**

Dr Jane A H Masoli ^73,^ ^91^.

**Metadata curation, and Samples and logistics:**

Dr Bridget A Knight ^73,^ ^91^, Dr Christopher R Jones ^73,^ ^91^, Mr Cherian Koshy ^1^, Miss Amy Ash ^1^, Dr Anna Casey ^71^, Dr Andrew Bosworth ^64,^ ^36^, Dr Liz Ratcliffe ^71^, Dr Li Xu-McCrae ^36^, Miss Hannah M Pymont ^64^, Ms Stephanie Hutchings ^64^, Dr Lisa Berry ^84^, Ms Katie Jones ^84^, Dr Fenella Halstead ^46^, Mr Thomas Davis ^21^, Dr Christopher Holmes ^16^, Prof Miren Iturriza-Gomara ^92^, Dr Anita O Lucaci ^92^, Dr Paul Anthony Randell ^38,^ ^104^, Dr Alison Cox ^38,^ ^104^, Pinglawathee Madona ^38,^ ^104^, Dr Kathryn Ann Harris ^30^, Dr Julianne Rose Brown ^30^, Dr Tabitha W Mahungu ^74^, Dr Dianne Irish-Tavares ^74^, Dr Tanzina Haque ^74^, Dr Jennifer Hart ^74^, Mr Eric Witele ^74^, Mrs Melisa Louise Fenton ^75^, Mr Steven Liggett ^79^, Dr Clive Graham ^56^, Ms Emma Swindells ^57^, Ms Jennifer Collins ^50^, Mr Gary Eltringham ^50^, Ms Sharon Campbell ^17^, Dr Patrick C McClure ^97^, Dr Gemma Clark ^15^, Dr Tim J Sloan ^60^, Mr Carl Jones ^15^ and Dr Jessica Lynch ^2,^ ^111^.

**Metadata curation, and Sequencing and analysis:**

Dr Ben Warne ^8^, Steven Leonard ^99^, Jillian Durham ^99^, Dr Thomas Williams ^90^, Dr Sam T Haldenby ^92^, Dr Nathaniel Storey ^30^, Dr Nabil-Fareed Alikhan ^70^, Dr Nadine Holmes ^18^, Dr Christopher Moore ^18^, Mr Matthew Carlile ^18^, Malorie Perry ^69^, Dr Noel Craine ^69^, Prof Ronan A Lyons ^80^, Miss Angela H Beckett ^13^, Salman Goudarzi ^77^, Christopher Fearn ^77^, Kate Cook ^77^, Hannah Dent ^77^ and Hannah Paul ^77^.

**Metadata curation, and Software and analysis tools:**

Robert Davies ^99^.

**Project administration, and Samples and logistics:**

Beth Blane ^88^, Sophia T Girgis ^88^, Dr Mathew A Beale ^99^, Katherine L Bellis ^99,^ ^88^, Matthew J Dorman ^99^, Eleanor Drury ^99^, Leanne Kane ^99^, Sally Kay ^99^, Dr Samantha McGuigan ^99^, Dr Rachel Nelson ^99^, Liam Prestwood ^99^, Dr Shavanthi Rajatileka ^99^, Dr Rahul Batra ^12^, Dr Rachel J Williams ^82^, Dr Mark Kristiansen ^82^, Dr Angie Green ^98^, Miss Anita Justice ^59^, Dr Adhyana I.K Mahanama ^81,^ ^102^ and Dr Buddhini Samaraweera ^81,^ ^102^.

**Project administration, and Sequencing and analysis:**

Dr Nazreen F Hadjirin ^88^ and Dr Joshua Quick ^41^.

**Project administration, and Software and analysis tools:**

Mr Radoslaw Poplawski ^41^.

**Samples and logistics, and Sequencing and analysis:**

Leanne M Kermack ^88^, Nicola Reynolds ^7^, Grant Hall ^24^, Yasmin Chaudhry ^24^, Malte L Pinckert ^24^, Dr Iliana Georgana ^24^, Dr Robin J Moll ^99^, Dr Alicia Thornton ^66^, Dr Richard Myers ^66^, Dr Joanne Stockton ^41^, Miss Charlotte A Williams ^82^, Dr Wen C Yew ^58^, Alexander J Trotter ^70^, Miss Amy Trebes ^98^, Mr George MacIntyre-Cockett ^98^, Alec Birchley ^69^, Alexander Adams ^69^, Amy Plimmer ^69^, Bree Gatica-Wilcox ^69^, Dr Caoimhe McKerr ^69^, Ember Hilvers ^69^, Hannah Jones ^69^, Dr Hibo Asad ^69^, Jason Coombes ^69^, Johnathan M Evans ^69^, Laia Fina ^69^, Lauren Gilbert ^69^, Lee Graham ^69^, Michelle Cronin ^69^, Sara Kumziene-SummerhaYes ^69^, Sarah Taylor ^69^, Sophie Jones ^69^, Miss Danielle C Groves ^93^, Mrs Peijun Zhang ^93^, Miss Marta Gallis ^93^ and Miss Stavroula F Louka ^93^.

**Samples and logistics, and Software and analysis tools:**

Dr Igor Starinskij ^48^.

**Sequencing and analysis, and Software and analysis tools:**

Dr Chris J Illingworth ^47^, Dr Chris Jackson ^47^, Ms Marina Gourtovaia ^99^, Gerry Tonkin-Hill ^99^, Kevin Lewis ^99^, Dr Jaime M Tovar-Corona ^99^, Dr Keith James ^99^, Dr Laura Baxter ^94^, Dr Mohammad T. Alam ^94^, Dr Richard J Orton ^48^, Dr Joseph Hughes ^48^, Dr Sreenu Vattipally ^48^, Dr Manon Ragonnet-Cronin ^39^, Dr Fabricia F. Nascimento ^39^, Mr David Jorgensen ^39^, Ms Olivia Boyd ^39^, Ms Lily Geidelberg ^39^, Dr Alex E Zarebski ^23^, Dr Jayna Raghwani ^23^, Dr Moritz UG Kraemer ^23^, Joel Southgate ^10,^ ^69^, Dr Benjamin B Lindsey ^93^ and Mr Timothy M Freeman ^93^.

**Software and analysis tools, and Visualisation:**

Jon-Paul Keatley ^99^, Dr Joshua B Singer ^48^, Leonardo de Oliveira Martins ^70^, Dr Corin A Yeats ^14^, Dr Khalil Abudahab ^14,^ ^114^, Mr Ben EW Taylor ^14,^ ^114^ and Mirko Menegazzo ^14^.

**Leadership and supervision:**

Prof John Danesh ^99^, Wendy Hogsden ^46^, Dr Sahar Eldirdiri ^21^, Mrs Anita Kenyon ^21^, Dr Jenifer Mason ^43^, Mr Trevor I Robinson ^43^, Prof Alison Holmes ^38,^ ^103^, Dr James Price ^38,^ ^103^, Prof John A Hartley ^82^, Dr Tanya Curran ^3^, Dr Alison E Mather ^70^, Dr Giri Shankar ^69^, Dr Rachel Jones ^69^, Dr Robin Howe ^69^ and Dr Sian Morgan ^9^.

**Metadata curation:**

Dr Elizabeth Wastenge ^53^, Dr Michael R Chapman ^34,^ ^88,^ ^99^, Mr Siddharth Mookerjee ^38,^ ^103^, Dr Rachael Stanley ^54^, Mrs Wendy Smith ^15^, Prof Timothy Peto ^59^, Dr David Eyre ^59^, Dr Derrick Crook ^59^, Dr Gabrielle Vernet ^33^, Dr Christine Kitchen ^10^, Huw Gulliver ^10^, Dr Ian Merrick ^10^, Prof Martyn Guest ^10^, Robert Munn ^10^, Dr Declan T Bradley ^63,^ ^72^, and Dr Tim Wyatt ^63^.

**Project administration:**

Dr Charlotte Beaver ^99^, Luke Foulser ^99^, Sophie Palmer ^88^, Carol M Churcher ^88^, Ellena Brooks ^88^, Kim S Smith ^88^, Dr Katerina Galai ^88^, Georgina M McManus ^88^, Dr Frances Bolt ^38,^ ^103^, Dr Francesc Coll ^19^, Lizzie Meadows ^70^, Dr Stephen W Attwood ^23^, Dr Alisha Davies ^69^, Elen De Lacy ^69^, Fatima Downing ^69^, Sue Edwards ^69^, Dr Garry P Scarlett ^76^, Mrs Sarah Jeremiah ^83^ and Dr Nikki Smith ^93^.

**Samples and logistics:**

Danielle Leek ^88^, Sushmita Sridhar ^88,^ ^99^, Sally Forrest ^88^, Claire Cormie ^88^, Harmeet K Gill ^88^, Joana Dias ^88^, Ellen E Higginson ^88^, Mailis Maes ^88^, Jamie Young ^88^, Michelle Wantoch ^7^, Sanger Covid Team (www.sanger.ac.uk/covid-team) ^99^, Dorota Jamrozy ^99^, Stephanie Lo ^99^, Dr Minal Patel ^99^, Verity Hill ^90^, Ms Claire M Bewshea ^91^, Prof Sian Ellard ^73,^ ^91^, Dr Cressida Auckland ^73^, Dr Ian Harrison ^66^, Dr Chloe Bishop ^66^, Dr Vicki Chalker ^66^, Dr Alex Richter ^85^, Dr Andrew Beggs ^85^, Dr Angus Best ^86^, Dr Benita Percival ^86^, Dr Jeremy Mirza ^86^, Dr Oliver Megram ^86^, Dr Megan Mayhew ^86^, Dr Liam Crawford ^86^, Dr Fiona Ashcroft ^86^, Dr Emma Moles-Garcia ^86^, Dr Nicola Cumley ^86^, Mr Richard Hopes ^64^, Dr Patawee Asamaphan ^48^, Mr Marc O Niebel ^48^, Prof Rory N Gunson ^100^, Dr Amanda Bradley ^52^, Dr Alasdair Maclean ^52^, Dr Guy Mollett ^52^, Dr Rachel Blacow ^52^, Mr Paul Bird ^16^, Mr Thomas Helmer ^16^, Miss Karlie Fallon ^16^, Dr Julian Tang ^16^, Dr Antony D Hale ^49^, Dr Louissa R Macfarlane-Smith ^49^, Katherine L Harper ^49^, Miss Holli Carden ^49^, Dr Nicholas W Machin ^45,^ ^64^, Ms Kathryn A Jackson ^92^, Dr Shazaad S Y Ahmad ^45,^ ^64^, Dr Ryan P George ^45^, Dr Lance Turtle ^92^, Mrs Elaine O'Toole ^43^, Mrs Joanne Watts ^43^, Mrs Cassie Breen ^43^, Mrs Angela Cowell ^43^, Ms Adela Alcolea-Medina ^32,^ ^96^, Ms Themoula Charalampous ^12,^ ^42^, Amita Patel ^11^, Dr Lisa J Levett ^35^, Dr Judith Heaney ^35^, Dr Aileen Rowan ^39^, Prof Graham P Taylor ^39^, Dr Divya Shah ^30^, Miss Laura Atkinson ^30^, Mr Jack CD Lee ^30^, Mr Adam P Westhorpe ^82^, Dr Riaz Jannoo ^82^, Dr Helen L Lowe ^82^, Miss Angeliki Karamani ^82^, Miss Leah Ensell ^82^, Mrs Wendy Chatterton ^35^, Miss Monika Pusok ^35^, Mrs Ashok Dadrah ^75^, Miss Amanda Symmonds ^75^, Dr Graciela Sluga ^44^, Dr Zoltan Molnar ^72^, Mr Paul Baker ^79^, Prof Stephen Bonner ^79^, Ms Sarah Essex ^79^, Dr Edward Barton ^56^, Ms Debra Padgett ^56^, Ms Garren Scott ^56^, Ms Jane Greenaway ^57^, Dr Brendan AI Payne ^50^, Dr Shirelle Burton-Fanning ^50^, Dr Sheila Waugh ^50^, Dr Veena Raviprakash ^17^, Ms Nicola Sheriff ^17^, Ms Victoria Blakey ^17^, ms Lesley-Anne Williams ^17^, Dr Jonathan Moore ^27^, Ms Susanne Stonehouse ^27^, Dr Louise Smith ^55^, Dr Rose K Davidson ^89^, Dr Luke Bedford ^26^, Dr Lindsay Coupland ^54^, Ms Victoria Wright ^18^, Dr Joseph G Chappell ^97^, Dr Theocharis Tsoleridis ^97^, Prof Jonathan Ball ^97^, Mrs Manjinder Khakh ^15^, Dr Vicki M Fleming ^15^, Dr Michelle M Lister ^15^, Dr Hannah C Howson-Wells ^15^, Dr Louise Berry ^15^, Dr Tim Boswell ^15^, Dr Amelia Joseph ^15^, Dr Iona Willingham ^15^, Dr Nichola Duckworth ^60^, Dr Sarah Walsh ^60^, Dr Emma Wise ^2,^ ^111^, Dr Nathan Moore ^2,^ ^111^, Miss Matilde Mori ^2,^ ^108,^ ^111^, Dr Nick Cortes ^2,^ ^111^, Dr Stephen Kidd ^2,^ ^111^, Dr Rebecca Williams ^33^, Laura Gifford ^69^, Miss Kelly Bicknell ^61^, Dr Sarah Wyllie ^61^, Miss Allyson Lloyd ^61^, Mr Robert Impey ^61^, Ms Cassandra S Malone ^6^, Mr Benjamin J Cogger ^6^, Nick Levene ^62^, Lynn Monaghan ^62^, Dr Alexander J Keeley ^93^, Dr David G Partridge ^78,^ ^93^, Dr Mohammad Raza ^78,^ ^93^, Dr Cariad Evans ^78,^ ^93^ and Dr Kate Johnson ^78,^ ^93^.

**Sequencing and analysis:**

Emma Betteridge ^99^, Ben W Farr ^99^, Scott Goodwin ^99^, Dr Michael A Quail ^99^, Carol Scott ^99^, Lesley Shirley ^99^, Scott AJ Thurston ^99^, Diana Rajan ^99^, Dr Iraad F Bronner ^99^, Louise Aigrain ^99^, Dr Nicholas M Redshaw ^99^, Dr Stefanie V Lensing ^99^, Shane McCarthy ^99^, Alex Makunin ^99^, Dr Carlos E Balcazar ^90^, Dr Michael D Gallagher ^90^, Dr Kathleen A Williamson ^90^, Thomas D Stanton ^90^, Ms Michelle L Michelsen ^91^, Ms Joanna Warwick-Dugdale ^91^, Dr Robin Manley ^91^, Ms Audrey Farbos ^91^, Dr James W Harrison ^91^, Dr Christine M Sambles ^91^, Dr David J Studholme ^91^, Dr Angie Lackenby ^66^, Dr Tamyo Mbisa ^66^, Dr Steven Platt ^66^, Mr Shahjahan Miah ^66^, Dr David Bibby ^66^, Dr Carmen Manso ^66^, Dr Jonathan Hubb ^66^, Dr Gavin Dabrera ^66^, Dr Mary Ramsay ^66^, Dr Daniel Bradshaw ^66^, Dr Ulf Schaefer ^66^, Dr Natalie Groves ^66^, Dr Eileen Gallagher ^66^, Dr David Lee ^66^, Dr David Williams ^66^, Dr Nicholas Ellaby ^66^, Hassan Hartman ^66^, Nikos Manesis ^66^, Vineet Patel ^66^, Juan Ledesma ^67^, Ms Katherine A Twohig ^67^, Dr Elias Allara ^64,^ ^88^, Ms Clare Pearson ^64,^ ^88^, Mr Jeffrey K. J. Cheng ^94^, Dr Hannah E. Bridgewater ^94^, Ms Lucy R. Frost ^94^, Ms Grace Taylor-Joyce ^94^, Dr Paul E Brown ^94^, Dr Lily Tong ^48^, Ms Alice Broos ^48^, Mr Daniel Mair ^48^, Mrs Jenna Nichols ^48^, Dr Stephen N Carmichael ^48^, Dr Katherine L Smollett ^40^, Dr Kyriaki Nomikou ^48^, Dr Elihu Aranday-Cortes ^48^, Ms Natasha Johnson ^48^, Dr Seema Nickbakhsh ^48,^ ^68^, Dr Edith E Vamos ^92^, Dr Margaret Hughes ^92^, Dr Lucille Rainbow ^92^, Mr Richard Eccles ^92^, Ms Charlotte Nelson ^92^, Dr Mark Whitehead ^92^, Dr Richard Gregory ^92^, Mr Matthew Gemmell ^92^, Ms Claudia Wierzbicki ^92^, Ms Hermione J Webster ^92^, Ms Chloe L Fisher ^28^, Mr Adrian W Signell ^20^, Dr Gilberto Betancor ^20^, Mr Harry D Wilson ^20^, Dr Gaia Nebbia ^12^, Dr Flavia Flaviani ^31^, Mr Alberto C Cerda ^96^, Ms Tammy V Merrill ^96^, Rebekah E Wilson ^96^, Mr Marius Cotic ^82^, Miss Nadua Bayzid ^82^, Dr Thomas Thompson ^72^, Dr Erwan Acheson ^72^, Prof Steven Rushton ^51^, Prof Sarah O'Brien ^51^, David J Baker ^70^, Steven Rudder ^70^, Alp Aydin ^70^, Dr Fei Sang ^18^, Dr Johnny Debebe ^18^, Dr Sarah Francois ^23^, Dr Tetyana I Vasylyeva ^23^, Dr Marina Escalera Zamudio ^23^, Mr Bernardo Gutierrez ^23^, Dr Angela Marchbank ^10^, Joshua Maksimovic ^9^, Karla Spellman ^9^, Kathryn McCluggage ^9^, Dr Mari Morgan ^69^, Robert Beer ^9^, Safiah Afifi ^9^, Trudy Workman ^10^, William Fuller ^10^, Catherine Bresner ^10^, Dr Adrienn Angyal ^93^, Dr Luke R Green ^93^, Dr Paul J Parsons ^93^, Miss Rachel M Tucker ^93^, Dr Rebecca Brown ^93^ and Mr Max Whiteley ^93^.

**Software and analysis tools:**

James Bonfield ^99^, Dr Christoph Puethe ^99^, Mr Andrew Whitwham ^99^, Jennifier Liddle ^99^, Dr Will Rowe ^41^, Dr Igor Siveroni ^39^, Dr Thanh Le-Viet ^70^ and Amy Gaskin ^69^.

**Visualisation:**

Dr Rob Johnson ^39^.

**1** Barking, Havering and Redbridge University Hospitals NHS Trust, **2** Basingstoke Hospital, **3** Belfast Health & Social Care Trust, **4** Betsi Cadwaladr University Health Board, **5** Big Data Institute, Nuffield Department of Medicine, University of Oxford, **6** Brighton and Sussex University Hospitals NHS Trust, **7** Cambridge Stem Cell Institute, University of Cambridge, **8** Cambridge University Hospitals NHS Foundation Trust, **9** Cardiff and Vale University Health Board, **10** Cardiff University, **11** Centre for Clinical Infection & Diagnostics Research, St. Thomas' Hospital and Kings College London, **12** Centre for Clinical Infection and Diagnostics Research, Department of Infectious Diseases, Guy's and St Thomas' NHS Foundation Trust, **13** Centre for Enzyme Innovation, University of Portsmouth (PORT), **14** Centre for Genomic Pathogen Surveillance, University of Oxford, **15** Clinical Microbiology Department, Queens Medical Centre, **16** Clinical Microbiology, University Hospitals of Leicester NHS Trust, **17** County Durham and Darlington NHS Foundation Trust, **18** Deep Seq, School of Life Sciences, Queens Medical Centre, University of Nottingham, **19** Department of Infection Biology, Faculty of Infectious & Tropical Diseases, London School of Hygiene & Tropical Medicine, **20** Department of Infectious Diseases, King's College London, **21** Department of Microbiology, Kettering General Hospital, **22** Departments of Infectious Diseases and Microbiology, Cambridge University Hospitals NHS Foundation Trust; Cambridge, UK, **23** Department of Zoology, University of Oxford, **24** Division of Virology, Department of Pathology, University of Cambridge, **25** East Kent Hospitals University NHS Foundation Trust, **26** East Suffolk and North Essex NHS Foundation Trust, **27** Gateshead Health NHS Foundation Trust, **28** Genomics Innovation Unit, Guy's and St. Thomas' NHS Foundation Trust, **29** Gloucestershire Hospitals NHS Foundation Trust, **30** Great Ormond Street Hospital for Children NHS Foundation Trust, **31** Guy's and St. Thomas’ BRC, **32** Guy's and St. Thomas’ Hospitals, **33** Hampshire Hospitals NHS Foundation Trust, **34** Health Data Research UK Cambridge, **35** Health Services Laboratories, **36** Heartlands Hospital, Birmingham, **37** Hub for Biotechnology in the Built Environment, Northumbria University, **38** Imperial College Hospitals NHS Trust, **39** Imperial College London, **40** Institute of Biodiversity, Animal Health & Comparative Medicine, **41** Institute of Microbiology and Infection, University of Birmingham, **42** King's College London, **43** Liverpool Clinical Laboratories, **44** Maidstone and Tunbridge Wells NHS Trust, **45** Manchester University NHS Foundation Trust, **46** Microbiology Department, Wye Valley NHS Trust, Hereford, **47** MRC Biostatistics Unit, University of Cambridge, **48** MRC-University of Glasgow Centre for Virus Research, **49** National Infection Service, PHE and Leeds Teaching Hospitals Trust, **50** Newcastle Hospitals NHS Foundation Trust, **51** Newcastle University, **52** NHS Greater Glasgow and Clyde, **53** NHS Lothian, **54** Norfolk and Norwich University Hospital, **55** Norfolk County Council, **56** North Cumbria Integrated Care NHS Foundation Trust, **57** North Tees and Hartlepool NHS Foundation Trust, **58** Northumbria University, **59** Oxford University Hospitals NHS Foundation Trust, **60** PathLinks, Northern Lincolnshire & Goole NHS Foundation Trust, **61** Portsmouth Hospitals University NHS Trust, **62** Princess Alexandra Hospital Microbiology Dept., **63** Public Health Agency, **64** Public Health England, **65** Public Health England, Clinical Microbiology and Public Health Laboratory, Cambridge, UK, **66** Public Health England, Colindale, **67** Public Health England, Colindale, **68** Public Health Scotland, **69** Public Health Wales NHS Trust, **70** Quadram Institute Bioscience, **71** Queen Elizabeth Hospital, **72** Queen's University Belfast, **73** Royal Devon and Exeter NHS Foundation Trust, **74** Royal Free NHS Trust, **75** Sandwell and West Birmingham NHS Trust, **76** School of Biological Sciences, University of Portsmouth (PORT), **77** School of Pharmacy and Biomedical Sciences, University of Portsmouth (PORT), **78** Sheffield Teaching Hospitals, **79** South Tees Hospitals NHS Foundation Trust, **80** Swansea University, **81** University Hospitals Southampton NHS Foundation Trust, **82** University College London, **83** University Hospital Southampton NHS Foundation Trust, **84** University Hospitals Coventry and Warwickshire, **85** University of Birmingham, **86** University of Birmingham Turnkey Laboratory, **87** University of Brighton, **88** University of Cambridge, **89** University of East Anglia, **90** University of Edinburgh, **91** University of Exeter, **92** University of Liverpool, **93** University of Sheffield, **94** University of Warwick, **95** University of Cambridge, **96** Viapath, Guy's and St Thomas' NHS Foundation Trust, and King's College Hospital NHS Foundation Trust, **97** Virology, School of Life Sciences, Queens Medical Centre, University of Nottingham, **98** Wellcome Centre for Human Genetics, Nuffield Department of Medicine, University of Oxford, **99** Wellcome Sanger Institute, **100** West of Scotland Specialist Virology Centre, NHS Greater Glasgow and Clyde, **101** Department of Medicine, University of Cambridge, **102** Ministry of Health, Sri Lanka, **103** NIHR Health Protection Research Unit in HCAI and AMR, Imperial College London, **104** North West London Pathology, **105** NU-OMICS, Northumbria University, **106** University of Kent, **107** University of Oxford, **108** University of Southampton, **109** University of Southampton School of Health Sciences, **110** University of Southampton School of Medicine, **111** University of Surrey, **112** Warwick Medical School and Institute of Precision Diagnostics, Pathology, UHCW NHS Trust, **113** Wellcome Africa Health Research Institute Durban and **114** Wellcome Genome Campus.
